# Comorbidities in autism spectrum disorder and their etiologies

**DOI:** 10.1101/2022.03.10.22272202

**Authors:** Vahe Khachadourian, Behrang Mahjani, Sven Sandin, Alexander Kolevzon, Joseph D. Buxbaum, Abraham Reichenberg, Magdalena Janecka

## Abstract

**Importance:** Perinatal exposures have been associated with autism spectrum disorder (ASD). It is unknown whether perinatal exposures are also associated with the burden of comorbidities in ASD across different medical domains.

**Objective:** To examine the burden and pattern of comorbidities in individuals with ASD, and evaluate the associations between perinatal exposures linked with ASD and distinct comorbidities.

**Design:** We used data from family-based study (SPARK) which recruited families with one or more children with a clinically confirmed ASD diagnosis across clinical sites throughout the US.

**Setting:** All data in SPARK are collected remotely to allow participants to complete the study protocol at their convenience.

**Participants:** To minimize recall bias of early-life exposures, we restricted the sample to individuals born between 1999 and 2019 who were less than 18 years of age at the time of registration into the study. The final analytic sample included 40,582 children with ASD and 11,389 non-ASD siblings.

**Exposures:** Perinatal exposures including preterm birth, prenatal infection, fetal alcohol syndrome, hypoxia at birth, bleeding into the brain during delivery, lead poisoning, brain infections, and traumatic brain injury.

**Main Outcomes and Measures:** Outcomes were based on parent report of professional diagnosis of neurological, cognitive, psychiatric, and physical disorders.

**Results:** Children with ASD had a substantially higher prevalence of all comorbidities compared to their siblings without an ASD diagnosis (all p-values <0.05). ADHD was the most common comorbidity, affecting 1 in every 3 children with ASD. In logistic regression models adjusted for covariates (annual household income, father’s and mother’s education, parental ages at childbirth, year of birth, age of the child at evaluation, race, and ethnicity), different exposures were associated with distinct patterns of comorbidities in ASD cases, including associations between preterm birth and difficulty gaining weight (OR=2.38; 95%CI=2.09-2.71) and traumatic brain injury and seizure or epilepsy (OR=4.75; 95%CI=3.25-6.95). We observed a similar pattern of associations in non-ASD siblings.

**Conclusions and Relevance:** Individuals with ASD experience a greater burden of perinatal exposures and comorbidities. The higher burden of comorbidities in this population could be partly attributable to the higher rates of perinatal exposures. Study findings, if replicated in other samples, can inform the etiology of comorbidity in ASD.

**KEY POINTS:** *Question:* Does the pattern of comorbidities in individuals with autism spectrum disorder vary by pre- and postnatal exposures?

*Findings:* The burden of pre- and postnatal exposures and comorbidity in children with autism spectrum disorder (ASD) is significantly higher than in their non-ASD siblings. Distinct pre- and postnatal exposures were associated with different patterns of comorbidities.

*Meaning:* Children with ASD experience a significantly higher burden of comorbidities which may arise due to distinct etiological factors.

## INTRODUCTION

Over the past 20 years, the prevalence of autism spectrum disorder (ASD) in the US has tripled, with current estimates indicating a prevalence of 1.85% among children^1^. The core ASD characteristics include social communication issues, restricted interests, and repetitive behaviors. In addition to the core diagnostic characteristics of ASD, affected individuals experience a higher burden of co-occurring conditions^2–9^ impacting a wide range of body systems. For instance, people with an ASD diagnosis are more likely to be overweight or obese^2,7,9^, or have a co-occurring accompanying psychiatric condition, such as anxiety and mood disorders^10^, compared to those without ASD. Prevalence estimates of several other conditions, including epilepsy^5^, sleep disorders^4^, and gastrointestinal disease^6^ are also much higher in ASD than those observed in the general population.

The comorbidities in ASD might have atypical manifestations and symptomatology, often making their recognition more difficult^11^. Despite these potential difficulties in the diagnosis, the reported comorbidity burden in ASD is still substantial - in a population-based study in Sweden, Lundström et al.^14^ found that over 50% of individuals with ASD had four or more comorbid conditions.

The observations of higher rates of certain health conditions in ASD^2–9^ suggest the possibility of a shared etiology between these different comorbidities and ASD. This possibility is substantiated by the observations that some ASD-associated risk genes often have pleiotropic effects on traits otherwise not seen in the majority of individuals with ASD (e.g., DDX3X and gait disturbance^15^). Despite this evidence, in non-syndromic forms of ASD, potential etiological overlaps between ASD and its comorbidities have not been systematically explored. This is despite the fact that investigation of comorbid conditions^16^ associated with ASD has the potential to offer insights into etiology.

The current study therefore aimed at addressing this gap by (1) describing the pattern of comorbidities in individuals with ASD, (2) examining if pre- and postnatal exposures previously associated with ASD may be linked to distinct comorbidities, thereby highlighting the potentially ‘pleiotropic’ effects of these exposures, and (3) conducting a comparative analysis in non-ASD siblings investigate the familial patterns of comorbidities in ASD, and examine if the effects of these environmental exposures on comorbidities might occur independently of the ASD diagnosis.

## METHODS

### Study design and population

We used the SPARK study database, launched by the Simons Foundation Autism Research Initiative (SFARI). The SPARK study recruited families with one or more children with an ASD diagnosis from 21 clinical sites throughout the US^17^. The SPARK database contains data collected from parents, including demographic characteristics, birth and developmental history, and medical diagnoses. All phenotypic data and biospecimens are collected remotely to allow participants to complete the study protocol at their convenience. The scientific community can access SPARK data free of charge.

We used all available SPARK datasets with phenotypic information, excluding the observations marked for possible unreliable ASD diagnosis or medical data. To minimize recall bias and increase the reliability of the data on early-life exposures, we restricted the sample to individuals born between 1999 and 2019 who were less than 18 years of age at the time of registration into the SPARK study. Finally, we excluded the individuals with a maternal age outside the range of 13-55 years or paternal age of less than 13 years at child’s birth, due to suspected administrative errors in the records.

The study protocol was reviewed by the institutional review board of the Icahn School of Medicine at Mount Sinai and received an exempt status.

### Exposure and outcome

Pregnancy and birth-related conditions served as the main exposure variables, including preterm birth (gestational age of <37 weeks), fetal alcohol syndrome, serious prenatal infection (e.g., German measles), hypoxia at birth, and bleeding into the brain during delivery. Hypoxia at birth was a binary variable that captured those children with hypoxia at birth who were admitted to a neonatal intensive care unit (NICU). Postnatal exposures included lead poisoning, brain infections such as bacterial meningitis, and encephalitis, and traumatic brain injury requiring hospitalization.

Medical history data in SPARK is based on parent report of professional diagnosis and is divided into multiple domains. A positive reply for each domain is followed by more specific and detailed questions regarding the medical conditions within that domain. In this study, as outcomes we considered medical conditions ascertained in the domains of a) birth or pregnancy complications, b) attention or behavior disorders, c) speech and language, intellectual disability/cognitive Impairment, learning disability (LD), or other developmental delay or developmental disability, d) growth conditions, e) neurological conditions, f) vision or hearing conditions, g) mood, depression, anxiety or obsessive-compulsive disorder (OCD), and h) sleep, feeding/eating or toileting problems.

For brevity and ease of reading, we refer to these outcomes as comorbidities, even when discussing the results from the non-ASD siblings, where unlike in children with ASD, there is no index condition.

### Covariates

Family and parental level covariates included the annual household income, and father’s and mother’s highest education levels, which served as a proxy for socio-economic status (SES), shown to be associated with ASD risk^18^, ASD diagnosis^19–21^ as well as health disparities^21,22^. Paternal and maternal ages at the time of delivery were adjusted to account for the possibility of higher rates of certain pre- and postnatal exposures and medical outcomes among the children of very young / older parents (e.g.,^23,24^). Child’s year of birth, age of the child at evaluation, and survey version, were additional covariates considered in this study, allowing us to control for the differences in the outcome/exposure rates that could be induced by the timing or the method of evaluation. Finally, we adjusted for child’s gender as well as the parent-reported race and ethnicity of the child to control for potential differences in the prevalence of the pre- and post-natal exposures^22,25^ and ASD diagnosis by gender, race and ethnicity^19–21,26^.

### Analysis

All analyses were performed using R programming language (version 4.0.0). Characteristics of the analytical sample by ASD status were summarized using means and frequencies. Prevalence of the exposures and comorbidities were estimated by ASD status. These estimates among the non-ASD siblings were standardized to the year of evaluation, age, and gender distribution observed among children with ASD in order to ensure that possible differences between these groups are not driven by demographic or recruitment characteristics. The standardization was performed by weighting the non-ASD siblings to mimic the year of evaluation, age, and gender distributions observed among children with ASD^27^. We repeated the analysis estimating the prevalence of exposures and comorbidities in sibling pairs (two siblings from each family) who were discordant for ASD diagnosis.

The binary logistic regression model was used to assess the associations between the exposures and outcomes of interest among children with ASD. Each outcome was evaluated in a separate model, including the exposures of interest and covariates. Considering potential clustering effects due to multiple deliveries among mothers in the study, we used clustered sandwich estimator, implemented in the clusterSEs package (v2.6.2). To avoid reporting unreliable associations due to the sparse data bias^28^, we did not analyze the exposure-outcome pairs with a recorded outcome frequency of less than 10 among exposed / unexposed children. To account for multiple testing, we applied false discovery rate correction^29^ to the empirical p-values.

Next, in each exposure-outcome model, we also adjusted for all other comorbidities grouped with the outcome (as per comorbidity groups identified by SPARK) in order to obtain estimates that are independent of the correlation patterns between these closely related comorbidities.

Finally, we estimated associations between the exposure-outcome pairs among the non-ASD siblings and contrasted the estimates with those obtained in the sample of children with ASD. Missing data for the covariates were imputed using *mice* package, applying multivariate imputation by chained equation^30^. The sample was restricted to the individuals with medical history data; hence for the exposure and outcome variables, no missing data imputation was required. We performed 5 imputations using information from all outcome and exposure variables as well as covariates (annual household income, and father’s and mother’s highest education levels, paternal age and maternal age, child’ gender and year of birth, age of the child at evaluation, survey version, race, and ethnicity).

## RESULTS

Among the individuals with complete medical data, there was a total of 42,564 individuals with ASD, and 11,390 of their non-ASD siblings (born 1999-2019). Forty-seven individuals with a maternal age beyond the range of 13-55 or paternal age of less than 13, and 1,936 individuals who were 18 and older at the time of registration into SPARK, were excluded from the analysis. The final analytic sample included 51,971 participants (40,582 with ASD and 11,389 siblings without ASD) born to 34,929 mothers.

Seventy-nine percent of children with ASD, and 49% non-ASD siblings were male. Mean maternal age at delivery was 29.5 among children with ASD and 29.1 among children without ASD diagnosis. Mean paternal age was also similar among cases and controls, 31.8 and 31.5 years respectively. Table 1 and Table S1 provide further details about sociodemographic characteristics of the analytical sample by ASD status.

**Table 1.** Demographic characteristics of the analytical sample by ASD status (N = 51,971)

|  | **ASD**<br>**(N = 40,582)** | **Control**<br>**(N = 11,389)** |
| --- | --- | --- |
| **Child's year of birth** |  |  |
| 1999–2004, n (%) | 5579 (13.7%) | 1638 (14.4%) |
| 2005–2009, n (%) | 11823 (29.1%) | 3481 (30.6%) |
| 2010–2014, n (%) | 15883 (39.1%) | 3985 (35.0%) |
| 2015–2019, n (%) | 7297 (18.0%) | 2285 (20.1%) |
| **Gender** |  |  |
| Female, n (%) | 8430 (20.8%) | 5805 (51.0%) |
| Male, n (%) | 32152 (79.2%) | 5584 (49.0%) |
| **Maternal age at delivery, mean (SD)** | 29.5 (6.0) | 29.1 (5.5) |
| **Paternal age at delivery, mean (SD)** | 31.8 (6.6) | 31.5 (6.2) |

### Prevalence of pre- and postnatal exposures

Individuals with ASD had a higher standardized prevalence of all of the pre-and postnatal exposures. The prevalence of intraventricular hemorrhage among children with ASD was 0.9% compared to 0.3% among non-ASD sibling controls (Figure 1). Similarly, prevalence of brain infection (0.3% vs <0.1%), fetal alcohol syndrome (1.2% vs 0.2%), infection in pregnancy (0.3% vs 0.1%), lead poisoning (0.3%, 0.1%), and traumatic brain injury (0.5%, 0.2%) were more than two-folds higher among children with ASD when compared to non-ASD sibling controls. Relative to non-ASD siblings, children with ASD diagnosis also had a higher prevalence of hypoxia at birth (6.9% vs 4.6%, p-value <0.05) and were more likely to be born preterm (13.2% vs 10.0%, p-value <0.05). The same pattern was observed when restricted the sample to pairs of siblings who were discordant for ASD diagnosis (Figure S1).

**Figure 1.**
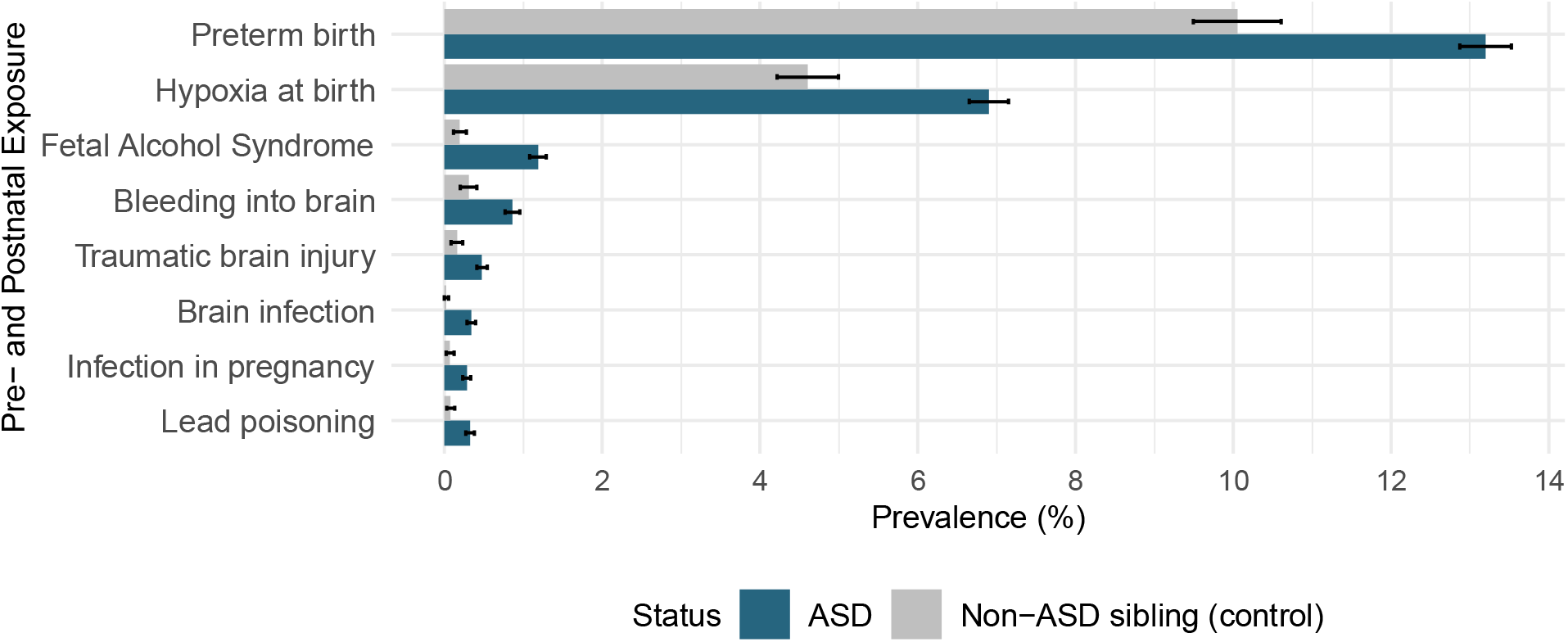
Prevalence of pre- and postnatal exposures by ASD status

### Prevalence and co-occurrence of comorbidities

Children with ASD had a substantially higher standardized prevalence of all comorbidities analyzed in this study compared to their siblings without an ASD diagnosis (all p-values <0.05). Attention deficit hyperactivity disorder (ADHD) was the most common comorbidity, affecting more 1 in every 3 children with ASD (35.3%), much higher than 1 in 6 (16.8%) among non-ASD siblings. Learning disability (23.5%) and intellectual disability (21.7%) were the next most-common comorbid conditions among children with ASD. Figure 2 presents prevalence of the analyzed comorbid conditions by ASD status and Figure S2 illustrates this in pairs of siblings who were discordant for ASD diagnosis.

**Figure 2.**
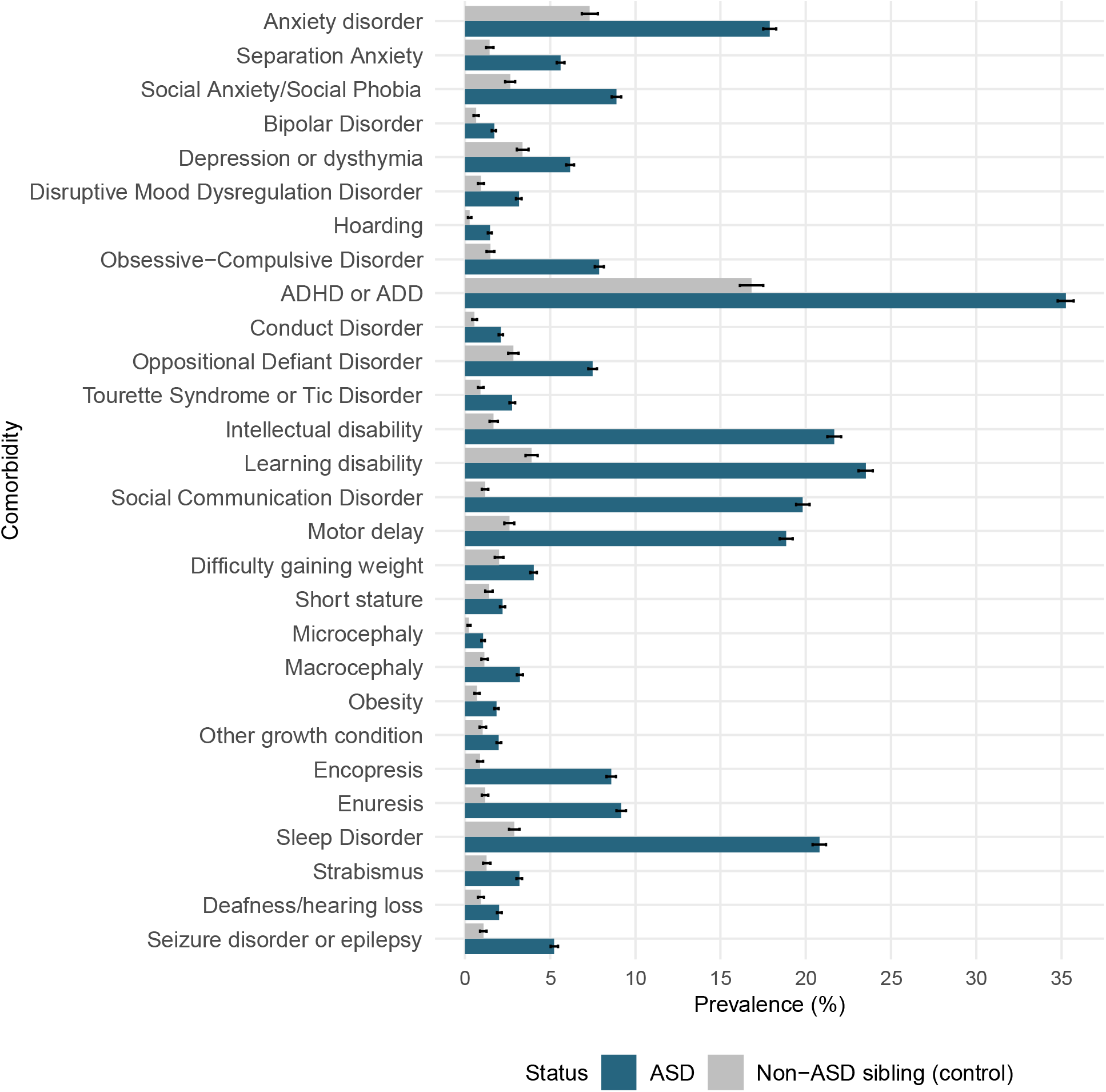
Prevalence of comorbid conditions by ASD status

The proportion of individuals with any comorbidity diagnosis was higher in individuals with ASD compared to their non-ASD siblings (Figure 3), mirroring the patterns observed for the individual diagnoses. Stratifying the sample based on the presence of at least one comorbidity, the burden of comorbidities remained higher among individuals with ASD than their non-ASD siblings. The correlations between pairs of the 28 comorbidities that served as outcomes in our analyses were predominantly positive (Figure S3). The correlations in the non-ASD siblings (Figure S4) were mostly similar to the ones observed in the ASD sample. However, the strength of the correlations in the non-ASD siblings was often lower than those observed in individuals with ASD.

**Figure 3.**
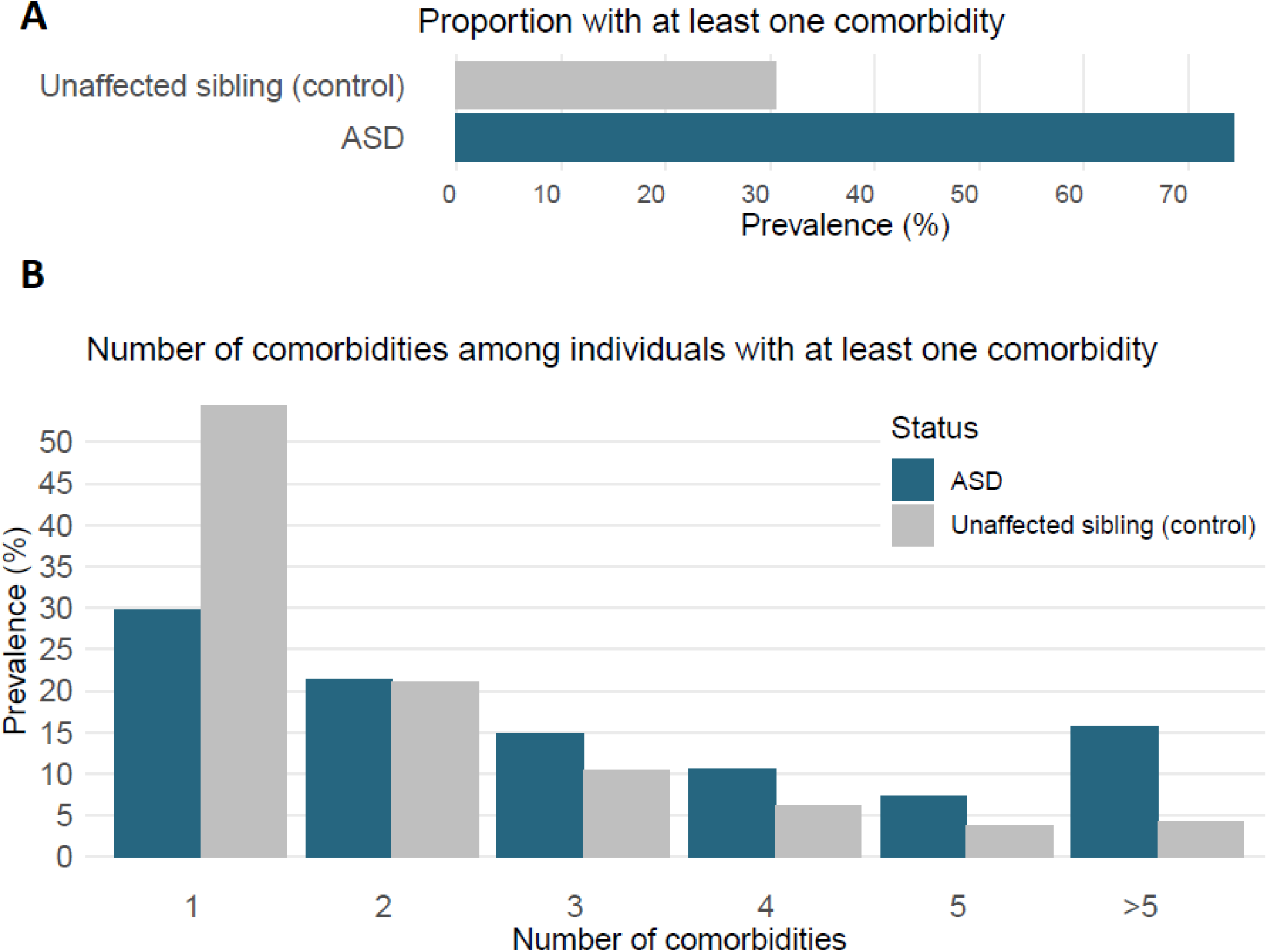
A: Prevalence of comorbidity by ASD status; B: Pattern of comorbidity among those with at least one comorbidity by ASD status

Comparing the prevalence of comorbidities between those children with no pre-and postnatal exposure to those with at least one pre-and postnatal exposure we found a relatively higher comorbidity prevalence among both ASD children as well as their non-ASD siblings (Figure S5).

### Association of pre-and postnatal exposures with comorbidity among children with ASD

There was a total of 224 possible exposure-comorbidity pairs, reflecting all combinations between 8 exposures and 28 outcomes (Figure 4a). Due to the rarity of some exposures, the association between 34 of these pairs could not be estimated among children with ASD. Among the remaining associations, we observed that exposure was associated with higher rates of comorbidity in 100 of these associations, and lower rates of comorbidity in 3, after applying a significance threshold of 0.05 to false discovery rate corrected p-values to adjust for multiple comparisons. Among the associations with a false discovery corrected p-value of ≥ 0.05, the point estimates (odds ratio) also often indicated increased rates of comorbidities associated with exposures, however, the wide confidence intervals for some of the estimates limited the scope for inference about these effects. The exposures associated with the highest number of comorbidities included preterm birth, hypoxia at birth, and fetal alcohol syndrome. Adjusting the associations for other comorbidities within the same cluster (related comorbidities), the magnitude of the odds ratios for some of the associations attenuated (Figure 4b). Although the number of significant associations reduced from 100 to 55, the overall pattern remained similar as in the previous analyses, with positive associations accounting for a large portion of the total significant associations. Tables 2S-9S provide frequency and associations of the exposure-outcome pairs.

**Figure 4.**
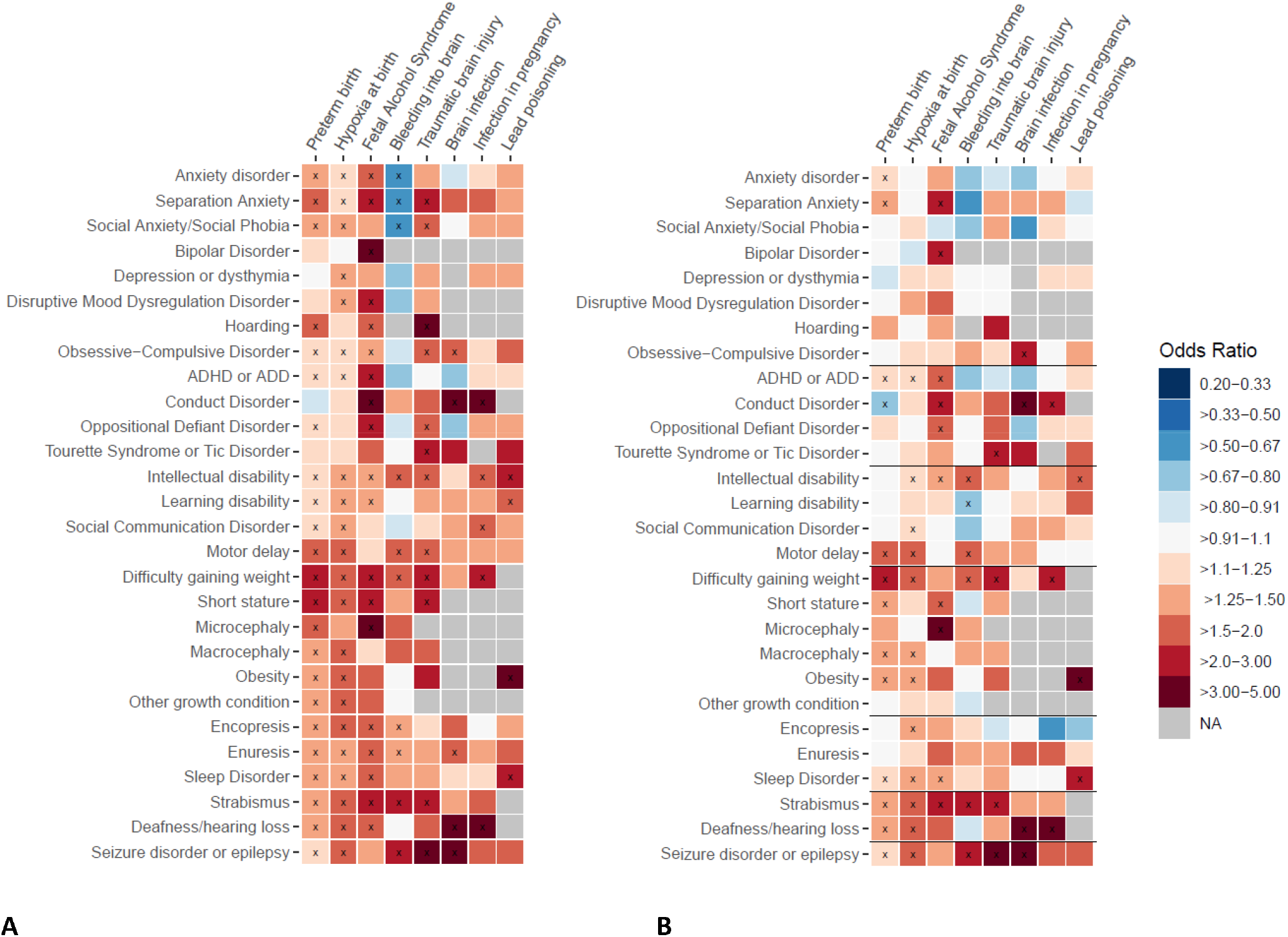
Associations between pre- and postnatal exposures and comorbidity in individuals with ASD ^x^: p-value < 0.05 **A:** The model for each comorbidity was adjusted for maternal and paternal education and age at delivery, child’s gender and year of birth, age of the child at evaluation, survey version, race, and annual household income. **B:** In addition to the covariates above, the model for each comorbidity was adjusted for other comorbidities within the same cluster (clusters are marked with solid black lines)

### Association of pre-and postnatal exposures with comorbidity among non-ASD siblings

To provide further context for interpretation of the results, we estimated the associations between exposures and ASD-comorbid conditions among the non-ASD siblings. Preterm birth and hypoxia at birth were the most common exposures in the non-ASD children allowing for evaluation of their associations with comorbidity within this smaller sample. Figure S6 presents these associations alongside the estimates obtained in the sample of individuals with ASD. Short stature, difficulty gaining weight, and motor delay had the strongest association with preterm birth, while motor delay, disruptive mood dysregulation disorder, and social communication disorder were the three comorbidities with the strongest association with hypoxia at birth. Most of the exposure-comorbidity estimates were suggestive of potential positive associations (OR > 1), however, for several of them, the width of the confidence intervals prevented drawing clear conclusions.

## DISCUSSION

We estimated the prevalence of pre- and postnatal exposures as well as comorbidity by ASD status, replicating the widely reported increased burden of health conditions among individuals with ASD. Our analyses show that the burden of comorbidity in ASD remains elevated even in comparison to the non-ASD siblings, suggesting that familial factors alone are unlikely to fully account for these observations.

Our results suggest that the pre- and postnatal exposures considered in this study are both (i) more common in children with ASD, compared to their non-ASD siblings, and (ii) associated with an increased burden of specific comorbidities — potentially indicating the ‘pleiotropic’ effects of these exposures (or their genetic underpinnings), on both ASD and its comorbidities (Figure 5). Furthermore, (iii) observing these exposure-comorbidity associations among the non-ASD siblings indicates that they may also occur independent of the ASD diagnosis. These results suggest that the higher rates of certain comorbidities in ASD may be partly attributable to the higher rates of the underlying risk factors (environmental exposures, or the underlying genetic variation) among the affected individuals, rather than be merely characteristic of ASD itself.

**Figure 5.**
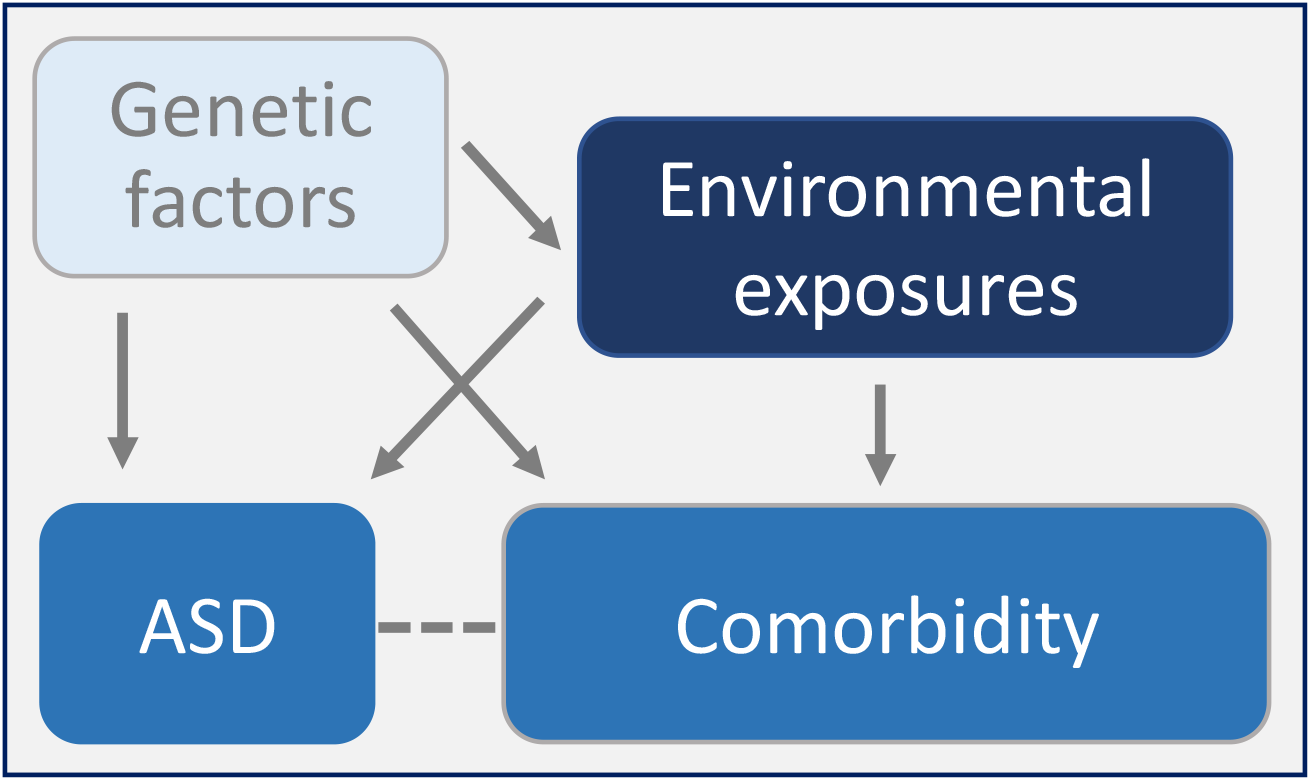
Potential relationships of genetic and environmental factors with ASD and comorbidity

In the initial analyses adjusting for covariates, we evaluated associations between 190 exposure-comorbidity pairs among individuals with ASD, observing predominantly positive associations. Further adjustment for other comorbid conditions potentially correlated with the outcome in each model decreased the number of significant associations, suggesting that some of the associations observed in the initial analyses were attributable to the clustering (positive correlation) of comorbid conditions. Nevertheless, even after this adjustment, the majority of the significant associations persisted, with hypoxia at birth, preterm birth, and fetal alcohol syndrome accounting for most of the significant exposure-comorbidity associations. Notably, these associations are in line with those reported in samples not ascertained for ASD diagnosis, including the association between strabismus and both pre-term birth^31^ and fetal alcohol syndrome^32^, and exposures directly affecting the brain (bleeding into brain, hypoxia at birth, brain infection, and traumatic brain injury) and epilepsy^33,34^. Although several of these associations were reported in previous studies conducted in the general population^31–34^, most of these associations were not evaluated or reported in a sample of individuals with ASD.

In addition to the systematic evaluation of these associations in individuals with ASD, we compared them with the associations observed in non-ASD siblings. While the rates of comorbidities were lower in siblings, many of the exposure-outcome associations remained significant in individuals without an ASD diagnosis, corroborating studies in the general population^31–34^ and indicating that the recorded associations were not attributable to the ASD diagnosis alone. This pattern of results, whereby exposures are seemingly independently associated with both ASD and its comorbidities, suggests their distinct effects on these different outcomes. Such ‘pleiotropic’ effects (underpinned, or not, by the genetic variation) could help explain the high frequency of comorbid diagnoses in ASD.

The study findings have several clinical and research implications. The heightened rates of co-occurring medical illnesses^9^ and health disparities^35^ among individuals with ASD, highlights the need for closer monitoring and better screening of these individuals for additional medical conditions. The positive correlations observed between certain exposures and comorbidities can potentially help with the early identification and treatment of these comorbidities. The exposure distribution and comorbidity pattern in individuals with ASD may also help explain the heterogeneity of ASD. Pre- and postnatal exposures in individuals with ASD were associated with distinct patterns of comorbidities, thus potentially offering a scope for stratification of these individuals for diagnosis and/or research purposes. These associations can provide further insights into the complex etiology of ASD and its accompanying comorbidities.

The study has several limitations. Given the non-probabilistic sampling and recruitment of study participants, we cannot exclude the risk of selection bias. Although the associations in our study are adjusted for a pool of covariates known to be correlated with pre- and postnatal exposures and/or comorbidities, no conclusions can be drawn about causality, especially given potential measurement error and uncontrolled confounding^36^ (e.g., access to care and genetic factors). Of note, even though adjusting for additional comorbidities yield exposure-comorbidity associations that are independent of other correlated comorbidities, such adjustments do not always result in estimates that are closer to the causal parameters. We also cannot rule out the possibility of measurement error (e.g., due to self-reporting, recall bias, or difficulty of diagnosing comorbidity in children with ASD) affecting the estimated associations.

Nevertheless, observing similar patterns as reported in other general population samples and the existence of several null and a few negative associations are suggestive that potential measurement error is unlikely to be a major concern.

A few of the covariates had high rates of missing values, nevertheless, we conducted multiple imputations, which, contingent on untestable assumptions regarding the missingness patterns, can eliminate missing data bias. The sensitivity analysis restricting the analytical sample to those with complete data yielded results that were consistent with those obtained from the analysis of the dataset with imputed missing covariates. Additionally, coding of some variables in SPARK does not distinguish between missing and “null” values, therefore, there is a chance that some of the observations coded as “null” or “no” were missing values instead. Given that the observed associations corroborate those observed in other, non-ASD samples, potential bias is minimal. Lack of data on the date of diagnosis of comorbidities limited our analytical approach and did not allow ‘time to event’ analysis in order to account for the fact that some of the individuals not yet diagnosed with ASD/comorbidity may receive a diagnosis later in life. Although SPARK is one of the largest ASD samples, the low prevalence of certain exposures and outcomes limited our statistical power in evaluating their associations, especially in the smaller sample of the non-ASD siblings.

This study followed a systematic approach, evaluating pre- and postnatal exposure and comorbidity associations in ASD. We demonstrated that the high burden of comorbidities in individuals with ASD are often associated with pre- and postnatal exposures also linked to ASD^37^. The higher prevalence of comorbidity, as well as higher number of comorbid conditions in individuals with ASD compared to their non-ASD siblings, draw attention to the importance of timely diagnosis and management of comorbidity in ASD, which in turn could offer improved prognosis and quality of life among these individuals. Finally, our preliminary evidence from associations between pre- and postnatal exposures and later comorbidities in the non-ASD siblings suggests that these associations are not limited to ASD and could partly be due to common underlying cause/s.

## Data Availability

The study data (SPARK study database) are available upon reasonable request to the Simons Foundation Autism Research Initiative (SFARI).

https://www.sfari.org/resource/spark/

## ACKNOWLEDGEMENTS

We would like to acknowledge the generous support of the Beatrice and Samuel A. Seaver Foundation. VK was supported by National Institute of Mental Health Award T32 MH122394. The content is solely the responsibility of the authors and does not necessarily represent the official views of the NIH or the authors’ employers.

## Notes

### Competing Interest Statement

The authors have declared no competing interest.

