## Supplemental Materials for "Comorbidities in autism spectrum disorder and their etiologies"

**Detailed Statistical Methods**

*Missing values*

Since the sample was restricted to those with medical history data, there were no missing values for exposure and outcome variables. In total 16,217 of children with ASD and 10909 of non-ASD siblings had missing information on one or more of the covariates. The missing data for the covariates were imputed using mice package, applying multivariate imputation by chained equation. Data for the exposure and outcome variables were complete and no missing data imputation was required. We performed 5 imputations using information from all outcome and exposure variables as well as covariates (annual household income, and father’s and mother’s highest education levels, paternal age and maternal age, gender, year of birth, age of the child at evaluation, survey version, race, and ethnicity) to impute missing information for covariates. The imputation model for each categorical covariates assumed multinomial logistic regression association. To test robustness of our findings from imputed covariates, we conducted complete case analysis and compared estimates of 56 exposure-outcome associations to the estimates obtained from the dataset with imputed missing covariates. The results from complete case analysis were consistent with those from the imputed dataset (Table S10-S11).

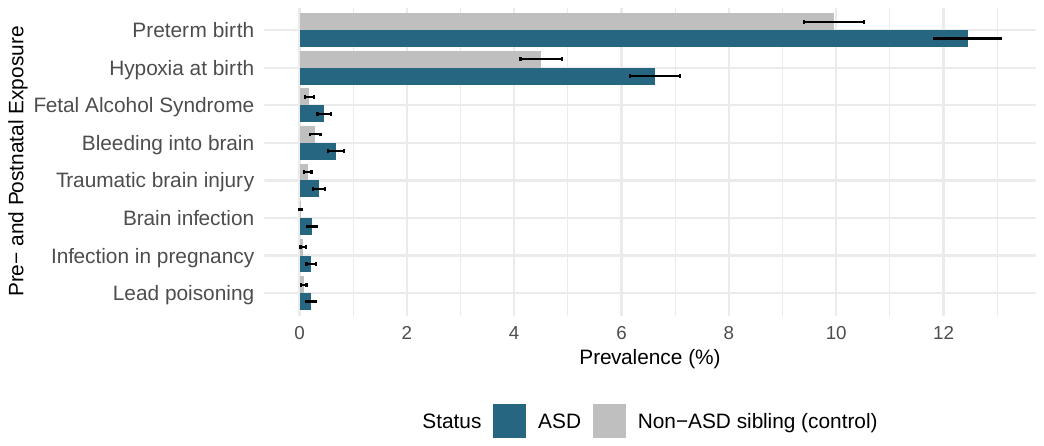

Figure S1 – Prevalence of pre- and postnatal exposures by ASD status, among pairs of siblings discordant for ASD diagnosis

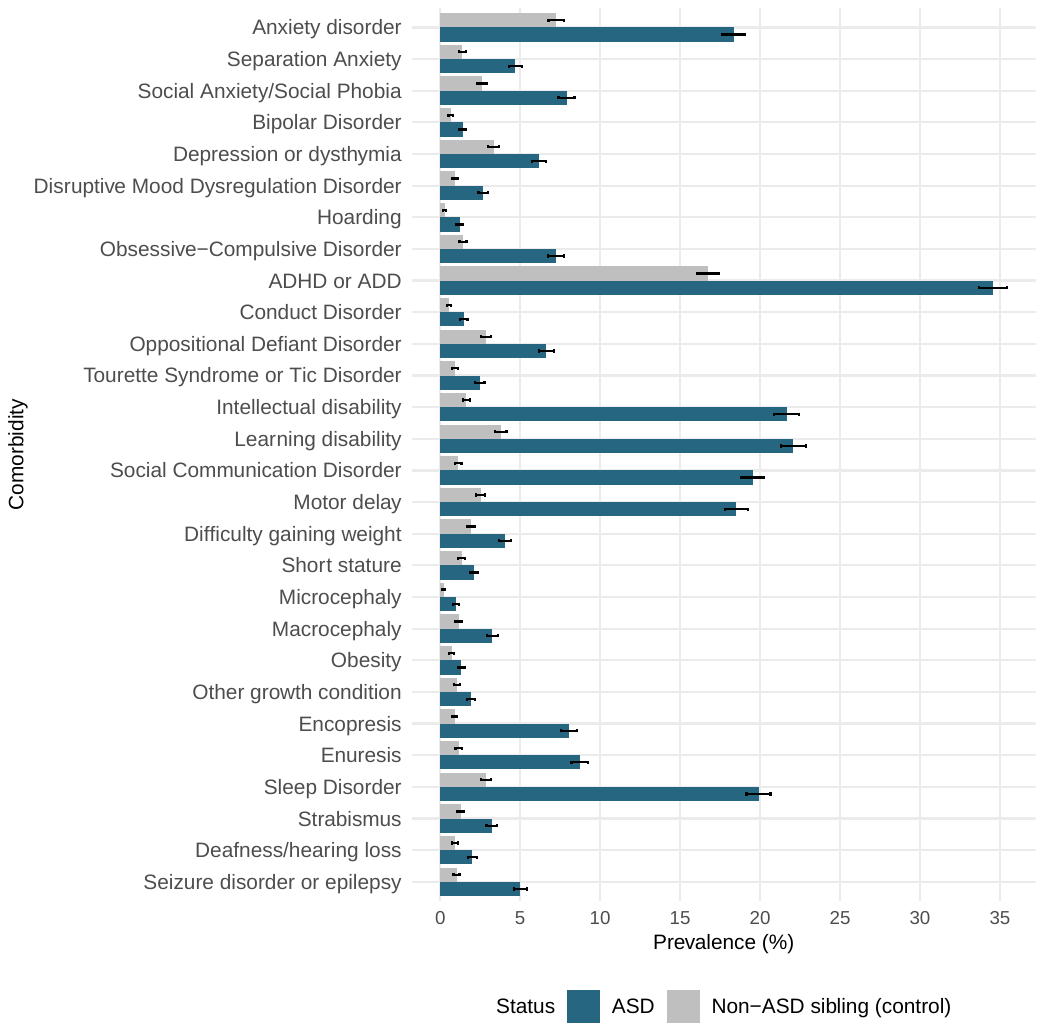

Figure S2 – Prevalence of comorbid conditions by ASD status, among pairs of siblings discordant for ASD diagnosis

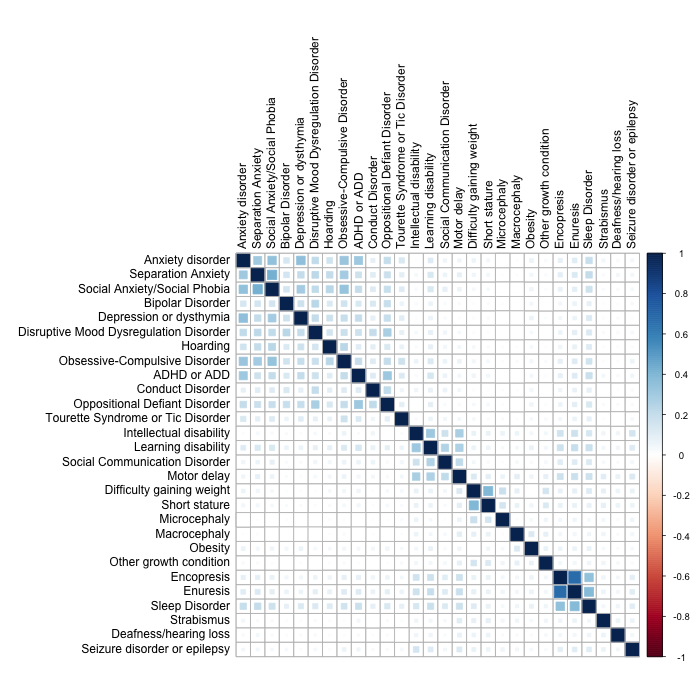

### Figure S3 – Correlation between various comorbid conditions in individuals with ASD diagnosis

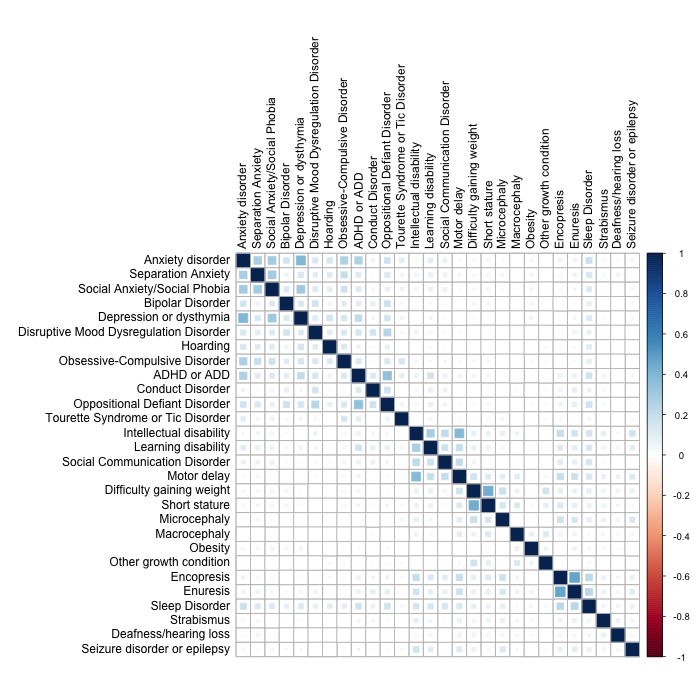

### Figure S4 – Correlation between various comorbid conditions in non-ASD siblings (controls)

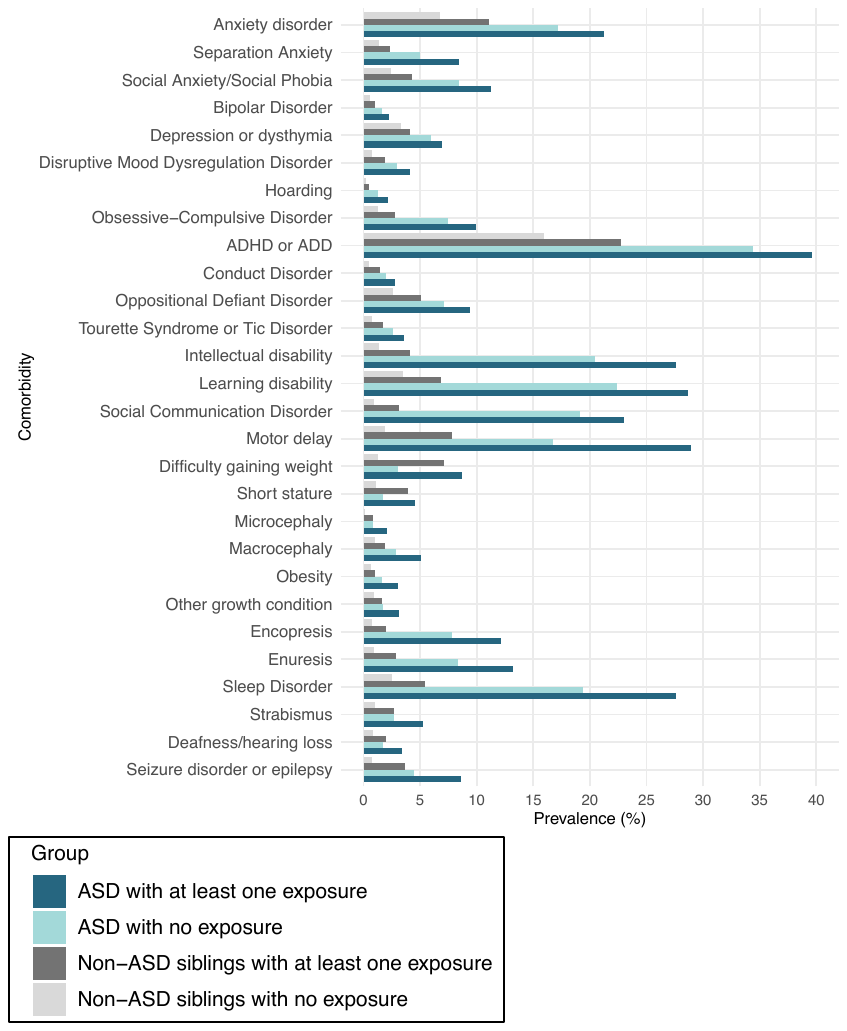

Figure S5 – Prevalence of comorbidities by ASD status and pre- and postnatal exposures

A) Preterm birth	B) Hypoxia at birth

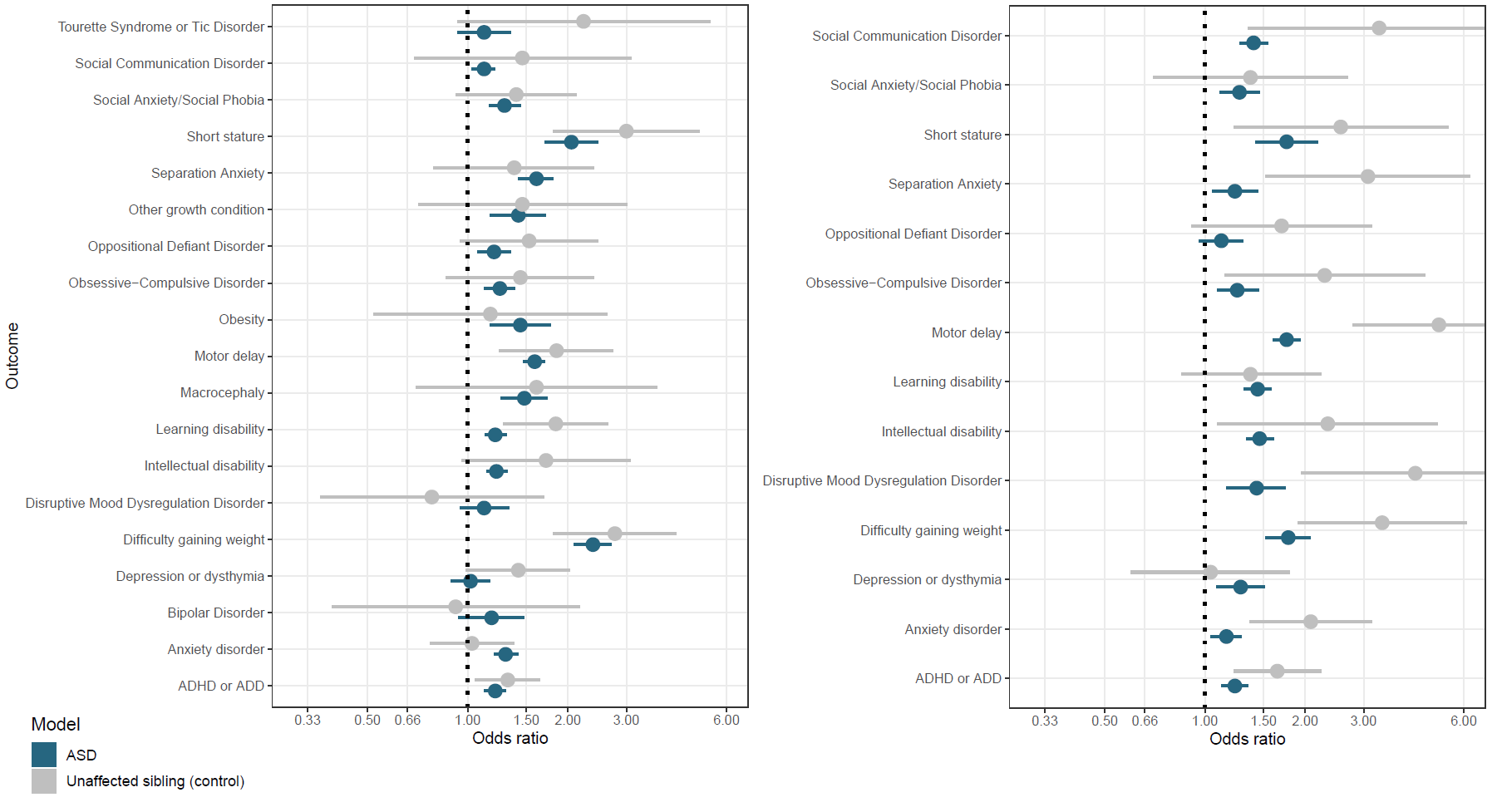

### Figure S6 - Associations between pre- and postnatal exposures and comorbidity by ASD status.

### The model for each comorbidity (outcome) was adjusted for maternal and paternal education and age at delivery, child’s gender and year of birth, age of the child at evaluation, survey version, race, and annual household income.

Table S1 – Additional demographic characteristics of the analytical sample by ASD status (N = 51,961)

|  | **ASD (N = 40,573)** | **Control (N = 11,388)** |
| --- | --- | --- |
| **Maternal education** |  |  |
| Associate degree | 5959 (14.7%) | 1333 (11.7%) |
| Bachelor’s degree | 9978 (24.6%) | 2436 (21.4%) |
| No high school | 115 (0.3%) | 596 (5.2%) |
| General education diploma | 1426 (3.5%) | 673 (5.9%) |
| Graduate/professional degree | 7211 (17.8%) | 1616 (14.2%) |
| High school graduate | 4327 (10.7%) | 1387 (12.2%) |
| Some college | 7224 (17.8%) | 1832 (16.1%) |
| Some high school | 1114 (2.7%) | 640 (5.6%) |
| Trade school | 3228 (8.0%) | 876 (7.7%) |
| **Paternal Education** |  |  |
| Associate degree | 3847 (9.5%) | 765 (6.7%) |
| Bachelor’s degree | 8243 (20.3%) | 2003 (17.6%) |
| No high school | 489 (1.2%) | 1027 (9.0%) |
| General education diploma | 2773 (6.8%) | 685 (6.0%) |
| Graduate/professional degree | 5740 (14.1%) | 1726 (15.2%) |
| High school graduate | 7989 (19.7%) | 2287 (20.1%) |
| Some college | 5762 (14.2%) | 1369 (12.0%) |
| Some high school | 2405 (5.9%) | 682 (6.0%) |
| Trade school | 3334 (8.2%) | 845 (7.4%) |
| **Annual household income in USD** |  |  |
| ≤ 20K | 5453 (13.4%) | 1546 (13.6%) |
| 21-35K | 6210 (15.3%) | 1870 (16.4%) |
| 36-50K | 5733 (14.1%) | 1402 (12.3%) |
| 51-65K | 4406 (10.9%) | 1113 (9.8%) |
| 66-80K | 4444 (11.0%) | 764 (6.7%) |
| 81-100K | 4554 (11.2%) | 1019 (8.9%) |
| 101-130K | 4362 (10.7%) | 1162 (10.2%) |
| 131-160K | 2013 (5.0%) | 848 (7.4%) |
| ≥161K | 3407 (8.4%) | 1665 (14.6%) |

Table S2 – Associations between preterm birth and comorbidity in individuals with ASD

| **Exposure** |  | **Exposed (n = 5356)** | **Unexposed (n = 35,226)** | **Model 1** | **Model 2** |
| --- | --- | --- | --- | --- | --- |
|  | **Outcome** | **n (%)^*^** | **n (%)^*^** | **OR [95% CI]** | **OR [95% CI]** |
| Preterm birth | Anxiety disorder | 1,169 (21.8%) | 6,090 (17.3%) | 1.30 [1.20-1.42] | 1.22 [1.11-1.34] |
|  | Separation Anxiety | 475 (8.9%) | 1,806 (5.1%) | 1.61 [1.42-1.81] | 1.45 [1.26-1.69] |
|  | Social Anxiety/Social Phobia | 622 (11.6%) | 2,986 (8.5%) | 1.29 [1.16-1.44] | 1.07 [0.94-1.21] |
|  | Bipolar Disorder | 116 (2.2%) | 577 (1.6%) | 1.18 [0.94-1.48] | 1.01 [0.79-1.31] |
|  | Depression or dysthymia | 369 (6.9%) | 2,134 (6.1%) | 1.02 [0.89-1.17] | 0.84 [0.72-0.98] |
|  | Disruptive Mood Dysregulation Disorder | 217 (4.1%) | 1,069 (3.0%) | 1.12 [0.95-1.33] | 0.92 [0.77-1.11] |
|  | Hoarding | 126 (2.4%) | 469 (1.3%) | 1.59 [1.27-1.98] | 1.30 [1.01-1.67] |
|  | Obsessive-Compulsive Disorder | 543 (10.1%) | 2,658 (7.5%) | 1.25 [1.12-1.39] | 1.03 [0.90-1.17] |
|  | ADHD or ADD | 2,151 (40.2%) | 12,156 (34.5%) | 1.21 [1.12-1.30] | 1.19 [1.10-1.28] |
|  | Conduct Disorder | 117 (2.2%) | 738 (2.1%) | 0.81 [0.65-1.02] | 0.73 [0.57-0.93] |
|  | Oppositional Defiant Disorder | 509 (9.5%) | 2,535 (7.2%) | 1.20 [1.07-1.35] | 1.15 [1.01-1.30] |
|  | Tourette Syndrome or Tic Disorder | 182 (3.4%) | 945 (2.7%) | 1.12 [0.93-1.35] | 1.08 [0.90-1.30] |
|  | Intellectual disability | 1,471 (27.5%) | 7,325 (20.8%) | 1.22 [1.14-1.32] | 1.07 [0.99-1.17] |
|  | Learning disability | 1,549 (28.9%) | 7,992 (22.7%) | 1.21 [1.13-1.31] | 1.07 [0.99-1.16] |
|  | Social Communication Disorder | 1,221 (22.8%) | 6,825 (19.4%) | 1.12 [1.03-1.21] | 0.98 [0.90-1.07] |
|  | Motor delay | 1,569 (29.3%) | 6,078 (17.3%) | 1.59 [1.47-1.71] | 1.54 [1.42-1.67] |
|  | Difficulty gaining weight | 504 (9.4%) | 1,132 (3.2%) | 2.38 [2.09-2.71] | 2.14 [1.85-2.48] |
|  | Short stature | 253 (4.7%) | 644 (1.8%) | 2.05 [1.71-2.46] | 1.34 [1.09-1.65] |
|  | Microcephaly | 116 (2.2%) | 316 (0.9%) | 1.92 [1.47-2.50] | 1.37 [1.03-1.82] |
|  | Macrocephaly | 275 (5.1%) | 1,034 (2.9%) | 1.48 [1.26-1.74] | 1.30 [1.10-1.53] |
|  | Obesity | 160 (3.0%) | 594 (1.7%) | 1.44 [1.17-1.78] | 1.34 [1.08-1.67] |
|  | Other growth condition | 163 (3.0%) | 644 (1.8%) | 1.42 [1.17-1.72] | 1.09 [0.88-1.34] |
|  | Encopresis | 631 (11.8%) | 2,853 (8.1%) | 1.26 [1.13-1.40] | 1.07 [0.92-1.26] |
|  | Enuresis | 679 (12.7%) | 3,044 (8.6%) | 1.28 [1.15-1.41] | 1.10 [0.94-1.29] |
|  | Sleep Disorder | 1,450 (27.1%) | 6,989 (19.8%) | 1.28 [1.18-1.38] | 1.22 [1.13-1.33] |
|  | Strabismus | 297 (5.5%) | 995 (2.8%) | 1.47 [1.27-1.71] | 1.45 [1.24-1.69] |
|  | Deafness/hearing loss | 180 (3.4%) | 634 (1.8%) | 1.48 [1.22-1.80] | 1.44 [1.18-1.75] |
|  | Seizure disorder or epilepsy | 426 (8.0%) | 1,698 (4.8%) | 1.25 [1.09-1.42] | 1.25 [1.09-1.42] |

* Percentages are column percentages
Model 1: The model for each comorbidity was adjusted for maternal and paternal education and age at delivery, child’s gender and year of birth, age of the child at evaluation, survey version, race, and annual household income.

Model 2: In addition to the covariates above, the model for each comorbidity was adjusted for other comorbidities with the same cluster (outcomes in the same cluster are highlighted in the same color)

Table S3 – Associations between hypoxia at birth and comorbidity in individuals with ASD

| **Exposure** |  | **Exposed (n = 2800)** | **Unexposed (n = 37,782)** | **Model 1** | **Model 2** |
| --- | --- | --- | --- | --- | --- |
|  | **Outcome** | **n (%)** | **n (%)** | **OR [95% CI]** | **OR [95% CI]** |
| Hypoxia at birth | Anxiety disorder | 614 (21.9%) | 6,645 (17.6%) | 1.16 [1.04-1.29] | 1.03 [0.91-1.18] |
|  | Separation Anxiety | 238 (8.5%) | 2,043 (5.4%) | 1.23 [1.05-1.44] | 1.05 [0.86-1.29] |
|  | Social Anxiety/Social Phobia | 343 (12.2%) | 3,265 (8.6%) | 1.27 [1.11-1.46] | 1.14 [0.97-1.35] |
|  | Bipolar Disorder | 59 (2.1%) | 634 (1.7%) | 1.10 [0.82-1.48] | 0.86 [0.60-1.22] |
|  | Depression or dysthymia | 224 (8.0%) | 2,279 (6.0%) | 1.28 [1.08-1.51] | 1.13 [0.93-1.36] |
|  | Disruptive Mood Dysregulation Disorder | 135 (4.8%) | 1,151 (3.0%) | 1.43 [1.16-1.75] | 1.31 [1.04-1.65] |
|  | Hoarding | 62 (2.2%) | 533 (1.4%) | 1.21 [0.89-1.67] | 0.99 [0.69-1.43] |
|  | Obsessive-Compulsive Disorder | 305 (10.9%) | 2,896 (7.7%) | 1.25 [1.09-1.45] | 1.14 [0.96-1.34] |
|  | ADHD or ADD | 1,164 (41.6%) | 13,143 (34.8%) | 1.23 [1.12-1.35] | 1.22 [1.10-1.34] |
|  | Conduct Disorder | 76 (2.7%) | 779 (2.1%) | 1.21 [0.91-1.62] | 1.16 [0.86-1.56] |
|  | Oppositional Defiant Disorder | 271 (9.7%) | 2,773 (7.3%) | 1.12 [0.96-1.30] | 1.01 [0.86-1.18] |
|  | Tourette Syndrome or Tic Disorder | 104 (3.7%) | 1,023 (2.7%) | 1.16 [0.92-1.47] | 1.12 [0.88-1.42] |
|  | Intellectual disability | 876 (31.3%) | 7,920 (21.0%) | 1.46 [1.33-1.61] | 1.20 [1.08-1.33] |
|  | Learning disability | 902 (32.2%) | 8,639 (22.9%) | 1.44 [1.31-1.58] | 1.13 [1.02-1.26] |
|  | Social Communication Disorder | 726 (25.9%) | 7,320 (19.4%) | 1.40 [1.27-1.55] | 1.16 [1.05-1.30] |
|  | Motor delay | 953 (34.0%) | 6,694 (17.7%) | 1.76 [1.60-1.94] | 1.56 [1.41-1.73] |
|  | Difficulty gaining weight | 294 (10.5%) | 1,342 (3.6%) | 1.78 [1.52-2.08] | 1.58 [1.32-1.90] |
|  | Short stature | 151 (5.4%) | 746 (2.0%) | 1.76 [1.42-2.19] | 1.20 [0.92-1.58] |
|  | Microcephaly | 62 (2.2%) | 370 (1.0%) | 1.43 [1.04-1.97] | 1.06 [0.75-1.49] |
|  | Macrocephaly | 168 (6.0%) | 1,141 (3.0%) | 1.54 [1.27-1.88] | 1.38 [1.13-1.69] |
|  | Obesity | 99 (3.5%) | 655 (1.7%) | 1.58 [1.23-2.03] | 1.49 [1.14-1.93] |
|  | Other growth condition | 99 (3.5%) | 708 (1.9%) | 1.56 [1.22-2.00] | 1.21 [0.93-1.58] |
|  | Encopresis | 391 (14.0%) | 3,093 (8.2%) | 1.51 [1.32-1.71] | 1.30 [1.07-1.57] |
|  | Enuresis | 409 (14.6%) | 3,314 (8.8%) | 1.45 [1.28-1.65] | 1.12 [0.92-1.36] |
|  | Sleep Disorder | 855 (30.5%) | 7,584 (20.1%) | 1.45 [1.31-1.59] | 1.34 [1.21-1.50] |
|  | Strabismus | 192 (6.9%) | 1,100 (2.9%) | 1.66 [1.38-2.00] | 1.61 [1.34-1.95] |
|  | Deafness/hearing loss | 117 (4.2%) | 697 (1.8%) | 1.72 [1.37-2.16] | 1.64 [1.30-2.08] |
|  | Seizure disorder or epilepsy | 302 (10.8%) | 1,822 (4.8%) | 1.83 [1.57-2.13] | 1.83 [1.57-2.13] |

* Percentages are column percentages
Model 1: The model for each comorbidity was adjusted for maternal and paternal education and age at delivery, child’s gender and year of birth, age of the child at evaluation, survey version, race, and annual household income.

Model 2: In addition to the covariates above, the model for each comorbidity was adjusted for other comorbidities with the same cluster (outcomes in the same cluster are highlighted in the same color)

Table S4 – Associations between fetal alcohol syndrome and comorbidity in individuals with ASD

| **Exposure** |  | **Exposed (n = 481)** | | **Unexposed (n = 40,101)** | **Model 1** | **Model 2** |
| --- | --- | --- | --- | --- | --- | --- |
|  | **Outcome** | **n (%)** | | **n (%)** | **OR [95% CI]** | **OR [95% CI]** |
| Fetal alcohol syndrome | Anxiety disorder | 123 (25.6%) | | 7,136 (17.8%) | 1.69 [1.31-2.18] | 1.41 [1.05-1.90] |
|  | Separation Anxiety | 78 (16.2%) | 2,203 (5.5%) | | 2.25 [1.69-3.01] | 2.14 [1.47-3.11] |
|  | Social Anxiety/Social Phobia | 69 (14.3%) | 3,539 (8.8%) | | 1.36 [1.00-1.85] | 0.83 [0.55-1.25] |
|  | Bipolar Disorder | 30 (6.2%) | 663 (1.7%) | | 3.07 [1.95-4.83] | 2.60 [1.56-4.31] |
|  | Depression or dysthymia | 43 (8.9%) | 2,460 (6.1%) | | 1.49 [1.04-2.13] | 1.22 [0.81-1.83] |
|  | Disruptive Mood Dysregulation Disorder | 42 (8.7%) | 1,244 (3.1%) | | 2.15 [1.49-3.11] | 1.60 [1.05-2.44] |
|  | Hoarding | 19 (4.0%) | 576 (1.4%) | | 1.93 [1.13-3.30] | 1.38 [0.68-2.79] |
|  | Obsessive-Compulsive Disorder | 60 (12.5%) | 3,141 (7.8%) | | 1.49 [1.10-2.01] | 1.11 [0.78-1.60] |
|  | ADHD or ADD | 251 (52.2%) | 14,056 (35.1%) | | 2.06 [1.66-2.56] | 1.69 [1.34-2.14] |
|  | Conduct Disorder | 48 (10.0%) | 807 (2.0%) | | 3.65 [2.51-5.30] | 2.47 [1.64-3.73] |
|  | Oppositional Defiant Disorder | 94 (19.5%) | 2,950 (7.4%) | | 2.69 [2.07-3.50] | 1.91 [1.45-2.52] |
|  | Tourette Syndrome or Tic Disorder | 23 (4.8%) | 1,104 (2.8%) | | 1.68 [1.07-2.64] | 1.30 [0.81-2.08] |
|  | Intellectual disability | 156 (32.4%) | 8,640 (21.5%) | | 1.47 [1.20-1.79] | 1.35 [1.08-1.68] |
|  | Learning disability | 171 (35.6%) | 9,370 (23.4%) | | 1.38 [1.12-1.70] | 1.24 [0.98-1.57] |
|  | Social Communication Disorder | 116 (24.1%) | 7,930 (19.8%) | | 1.15 [0.91-1.44] | 1.01 [0.80-1.29] |
|  | Motor delay | 133 (27.7%) | 7,514 (18.7%) | | 1.21 [0.97-1.50] | 1.02 [0.81-1.30] |
|  | Difficulty gaining weight | 53 (11.0%) | 1,583 (3.9%) | | 2.11 [1.53-2.93] | 1.34 [0.88-2.05] |
|  | Short stature | 39 (8.1%) | 858 (2.1%) | | 2.65 [1.82-3.87] | 1.82 [1.15-2.88] |
|  | Microcephaly | 25 (5.2%) | 407 (1.0%) | | 4.17 [2.71-6.43] | 3.14 [1.89-5.22] |
|  | Macrocephaly | 19 (4.0%) | 1,290 (3.2%) | | 1.12 [0.68-1.82] | 0.95 [0.56-1.60] |
|  | Obesity | 19 (4.0%) | 735 (1.8%) | | 1.76 [1.04-2.98] | 1.72 [1.00-2.96] |
|  | Other growth condition | 18 (3.7%) | 789 (2.0%) | | 1.67 [1.02-2.72] | 1.17 [0.68-2.03] |
|  | Encopresis | 83 (17.3%) | 3,401 (8.5%) | | 1.80 [1.38-2.35] | 1.26 [0.81-1.98] |
|  | Enuresis | 92 (19.1%) | 3,631 (9.1%) | | 1.89 [1.46-2.44] | 1.52 [1.01-2.28] |
|  | Sleep Disorder | 170 (35.3%) | 8,269 (20.6%) | | 1.63 [1.32-2.02] | 1.39 [1.10-1.76] |
|  | Strabismus | 40 (8.3%) | 1,252 (3.1%) | | 2.20 [1.57-3.08] | 2.14 [1.52-3.03] |
|  | Deafness/hearing loss | 22 (4.6%) | 792 (2.0%) | | 1.84 [1.16-2.91] | 1.68 [1.05-2.70] |
|  | Seizure disorder or epilepsy | 37 (7.7%) | 2,087 (5.2%) | | 1.28 [0.89-1.83] | 1.28 [0.89-1.83] |

* Percentages are column percentages
Model 1: The model for each comorbidity was adjusted for maternal and paternal education and age at delivery, child’s gender and year of birth, age of the child at evaluation, survey version, race, and annual household income.

Model 2: In addition to the covariates above, the model for each comorbidity was adjusted for other comorbidities with the same cluster (outcomes in the same cluster are highlighted in the same color)

Table S5 – Associations between bleeding into brain and comorbidity in individuals with ASD

| **Exposure** |  | **Exposed (n = 349)** | | **Unexposed (n = 40,233)** | **Model 1** | **Model 2** |
| --- | --- | --- | --- | --- | --- | --- |
|  | **Outcome** | **n (%)** | | **n (%)** | **OR [95% CI]** | **OR [95% CI]** |
| Bleeding into brain | Anxiety disorder | 59 (16.9%) | | 7,200 (17.9%) | 0.63 [0.46-0.86] | 0.71 [0.50-1.01] |
|  | Separation Anxiety | 22 (6.3%) | 2,259 (5.6%) | | 0.54 [0.34-0.86] | 0.65 [0.36-1.18] |
|  | Social Anxiety/Social Phobia | 30 (8.6%) | 3,578 (8.9%) | | 0.59 [0.40-0.86] | 0.75 [0.47-1.21] |
|  | Bipolar Disorder | 3 (0.9%) | 690 (1.7%) | | NA | NA |
|  | Depression or dysthymia | 20 (5.7%) | 2,483 (6.2%) | | 0.71 [0.44-1.15] | 1.03 [0.60-1.76] |
|  | Disruptive Mood Dysregulation Disorder | 13 (3.7%) | 1,273 (3.2%) | | 0.73 [0.41-1.31] | 1.06 [0.55-2.05] |
|  | Hoarding | 4 (1.1%) | 591 (1.5%) | | NA | NA |
|  | Obsessive-Compulsive Disorder | 36 (10.3%) | 3,165 (7.9%) | | 0.87 [0.60-1.28] | 1.32 [0.88-2.00] |
|  | ADHD or ADD | 134 (38.4%) | 14,173 (35.2%) | | 0.80 [0.63-1.03] | 0.80 [0.61-1.04] |
|  | Conduct Disorder | 13 (3.7%) | 842 (2.1%) | | 1.29 [0.64-2.57] | 1.50 [0.74-3.03] |
|  | Oppositional Defiant Disorder | 32 (9.2%) | 3,012 (7.5%) | | 0.83 [0.55-1.26] | 0.95 [0.61-1.47] |
|  | Tourette Syndrome or Tic Disorder | 15 (4.3%) | 1,112 (2.8%) | | 0.97 [0.55-1.71] | 1.06 [0.60-1.88] |
|  | Intellectual disability | 145 (41.5%) | 8,651 (21.5%) | | 1.67 [1.33-2.10] | 1.61 [1.25-2.08] |
|  | Learning disability | 108 (30.9%) | 9,433 (23.4%) | | 0.95 [0.74-1.22] | 0.70 [0.53-0.92] |
|  | Social Communication Disorder | 83 (23.8%) | 7,963 (19.8%) | | 0.88 [0.68-1.16] | 0.73 [0.55-0.98] |
|  | Motor delay | 160 (45.8%) | 7,487 (18.6%) | | 1.86 [1.47-2.36] | 1.90 [1.48-2.44] |
|  | Difficulty gaining weight | 63 (18.1%) | 1,573 (3.9%) | | 1.71 [1.25-2.34] | 1.66 [1.14-2.41] |
|  | Short stature | 26 (7.4%) | 871 (2.2%) | | 1.30 [0.81-2.09] | 0.84 [0.45-1.55] |
|  | Microcephaly | 15 (4.3%) | 417 (1.0%) | | 1.63 [0.90-2.97] | 1.41 [0.72-2.76] |
|  | Macrocephaly | 29 (8.3%) | 1,280 (3.2%) | | 1.53 [0.99-2.37] | 1.42 [0.89-2.25] |
|  | Obesity | 12 (3.4%) | 742 (1.8%) | | 0.97 [0.49-1.91] | 0.98 [0.50-1.90] |
|  | Other growth condition | 13 (3.7%) | 794 (2.0%) | | 1.09 [0.57-2.07] | 0.82 [0.42-1.59] |
|  | Encopresis | 64 (18.3%) | 3,420 (8.5%) | | 1.46 [1.07-1.99] | 1.25 [0.80-1.95] |
|  | Enuresis | 70 (20.1%) | 3,653 (9.1%) | | 1.46 [1.08-1.97] | 1.30 [0.85-1.99] |
|  | Sleep Disorder | 122 (35.0%) | 8,317 (20.7%) | | 1.29 [1.01-1.65] | 1.13 [0.83-1.53] |
|  | Strabismus | 55 (15.8%) | 1,237 (3.1%) | | 2.74 [1.96-3.82] | 2.75 [1.96-3.87] |
|  | Deafness/hearing loss | 16 (4.6%) | 798 (2.0%) | | 1.10 [0.64-1.92] | 0.87 [0.48-1.55] |
|  | Seizure disorder or epilepsy | 75 (21.5%) | 2,049 (5.1%) | | 2.36 [1.72-3.24] | 2.36 [1.72-3.24] |

* Percentages are column percentages
Model 1: The model for each comorbidity was adjusted for maternal and paternal education and age at delivery, child’s gender and year of birth, age of the child at evaluation, survey version, race, and annual household income.

Model 2: In addition to the covariates above, the model for each comorbidity was adjusted for other comorbidities with the same cluster (outcomes in the same cluster are highlighted in the same color)

Table S6 – Associations between traumatic brain injury and comorbidity in individuals with ASD

| **Exposure** |  | **Exposed (n = 193)** | | **Unexposed (n = 40,389)** | **Model 1** | **Model 2** |
| --- | --- | --- | --- | --- | --- | --- |
|  | **Outcome** | **n (%)** | | **n (%)** | **OR [95% CI]** | **OR [95% CI]** |
| Traumatic brain injury | Anxiety disorder | 51 (26.4%) | | 7,208 (17.8%) | 1.26 [0.89-1.80] | 0.87 [0.53-1.41] |
|  | Separation Anxiety | 29 (15.0%) | 2,252 (5.6%) | | 2.07 [1.34-3.20] | 1.45 [0.85-2.46] |
|  | Social Anxiety/Social Phobia | 38 (19.7%) | 3,570 (8.8%) | | 1.93 [1.32-2.81] | 1.26 [0.74-2.13] |
|  | Bipolar Disorder | 9 (4.7%) | 684 (1.7%) | | NA | NA |
|  | Depression or dysthymia | 23 (11.9%) | 2,480 (6.1%) | | 1.50 [0.94-2.41] | 1.00 [0.57-1.77] |
|  | Disruptive Mood Dysregulation Disorder | 14 (7.3%) | 1,272 (3.1%) | | 1.50 [0.85-2.65] | 0.99 [0.55-1.76] |
|  | Hoarding | 13 (6.7%) | 582 (1.4%) | | 3.39 [1.87-6.18] | 2.22 [1.00-4.93] |
|  | Obsessive-Compulsive Disorder | 34 (17.6%) | 3,167 (7.8%) | | 1.79 [1.20-2.66] | 1.21 [0.68-2.14] |
|  | ADHD or ADD | 84 (43.5%) | 14,223 (35.2%) | | 1.07 [0.79-1.45] | 0.89 [0.63-1.25] |
|  | Conduct Disorder | 12 (6.2%) | 843 (2.1%) | | 1.80 [0.90-3.60] | 1.54 [0.78-3.07] |
|  | Oppositional Defiant Disorder | 32 (16.6%) | 3,012 (7.5%) | | 1.82 [1.19-2.77] | 1.62 [1.00-2.62] |
|  | Tourette Syndrome or Tic Disorder | 20 (10.4%) | 1,107 (2.7%) | | 2.89 [1.73-4.84] | 2.59 [1.53-4.39] |
|  | Intellectual disability | 77 (39.9%) | 8,719 (21.6%) | | 1.72 [1.27-2.32] | 1.47 [1.07-2.02] |
|  | Learning disability | 71 (36.8%) | 9,470 (23.4%) | | 1.36 [1.00-1.87] | 1.07 [0.78-1.49] |
|  | Social Communication Disorder | 50 (25.9%) | 7,996 (19.8%) | | 1.22 [0.87-1.71] | 0.99 [0.69-1.42] |
|  | Motor delay | 76 (39.4%) | 7,571 (18.7%) | | 1.75 [1.27-2.41] | 1.48 [1.02-2.15] |
|  | Difficulty gaining weight | 32 (16.6%) | 1,604 (4.0%) | | 2.36 [1.54-3.61] | 2.26 [1.32-3.88] |
|  | Short stature | 16 (8.3%) | 881 (2.2%) | | 2.10 [1.16-3.81] | 1.30 [0.60-2.85] |
|  | Microcephaly | 7 (3.6%) | 425 (1.1%) | | NA | NA |
|  | Macrocephaly | 14 (7.3%) | 1,295 (3.2%) | | 1.66 [0.91-3.04] | 1.40 [0.73-2.68] |
|  | Obesity | 11 (5.7%) | 743 (1.8%) | | 2.02 [1.00-4.06] | 1.69 [0.76-3.77] |
|  | Other growth condition | 8 (4.1%) | 799 (2.0%) | | NA | NA |
|  | Encopresis | 29 (15.0%) | 3,455 (8.6%) | | 1.15 [0.73-1.81] | 0.82 [0.40-1.66] |
|  | Enuresis | 36 (18.7%) | 3,687 (9.1%) | | 1.36 [0.89-2.10] | 1.40 [0.77-2.57] |
|  | Sleep Disorder | 67 (34.7%) | 8,372 (20.7%) | | 1.38 [0.99-1.91] | 1.31 [0.92-1.88] |
|  | Strabismus | 28 (14.5%) | 1,264 (3.1%) | | 2.47 [1.54-3.94] | 2.37 [1.46-3.85] |
|  | Deafness/hearing loss | 11 (5.7%) | 803 (2.0%) | | 1.65 [0.86-3.19] | 1.46 [0.74-2.87] |
|  | Seizure disorder or epilepsy | 62 (32.1%) | 2,062 (5.1%) | | 4.75 [3.25-6.95] | 4.75 [3.25-6.95] |

* Percentages are column percentages
Model 1: The model for each comorbidity was adjusted for maternal and paternal education and age at delivery, child’s gender and year of birth, age of the child at evaluation, survey version, race, and annual household income.

Model 2: In addition to the covariates above, the model for each comorbidity was adjusted for other comorbidities with the same cluster (outcomes in the same cluster are highlighted in the same color)

Table S7 – Associations between brain infection and comorbidity in individuals with ASD

| **Exposure** |  | **Exposed (n = 138)** | | **Unexposed (n = 40,444)** | **Model 1** | **Model 2** |
| --- | --- | --- | --- | --- | --- | --- |
|  | **Outcome** | **n (%)** | | **n (%)** | **OR [95% CI]** | **OR [95% CI]** |
| Brain infection | Anxiety disorder | 29 (21.0%) | | 7,230 (17.9%) | 0.89 [0.56-1.41] | 0.77 [0.45-1.30] |
|  | Separation Anxiety | 16 (11.6%) | 2,265 (5.6%) | | 1.64 [0.85-3.17] | 1.41 [0.50-4.03] |
|  | Social Anxiety/Social Phobia | 17 (12.3%) | 3,591 (8.9%) | | 1.01 [0.58-1.76] | 0.64 [0.28-1.49] |
|  | Bipolar Disorder | 2 (1.4%) | 691 (1.7%) | | NA | NA |
|  | Depression or dysthymia | 8 (5.8%) | 2,495 (6.2%) | | NA | NA |
|  | Disruptive Mood Dysregulation Disorder | 8 (5.8%) | 1,278 (3.2%) | | NA | NA |
|  | Hoarding | 5 (3.6%) | 590 (1.5%) | | NA | NA |
|  | Obsessive-Compulsive Disorder | 24 (17.4%) | 3,177 (7.9%) | | 1.93 [1.14-3.27] | 2.19 [1.23-3.90] |
|  | ADHD or ADD | 48 (34.8%) | 14,259 (35.3%) | | 0.76 [0.51-1.13] | 0.71 [0.46-1.09] |
|  | Conduct Disorder | 12 (8.7%) | 843 (2.1%) | | 3.91 [2.07-7.40] | 4.85 [2.47-9.50] |
|  | Oppositional Defiant Disorder | 11 (8.0%) | 3,033 (7.5%) | | 0.80 [0.42-1.54] | 0.71 [0.35-1.45] |
|  | Tourette Syndrome or Tic Disorder | 10 (7.2%) | 1,117 (2.8%) | | 2.05 [1.02-4.11] | 2.20 [1.06-4.55] |
|  | Intellectual disability | 40 (29.0%) | 8,756 (21.6%) | | 1.22 [0.83-1.79] | 1.01 [0.67-1.52] |
|  | Learning disability | 48 (34.8%) | 9,493 (23.5%) | | 1.37 [0.94-2.01] | 1.20 [0.80-1.79] |
|  | Social Communication Disorder | 37 (26.8%) | 8,009 (19.8%) | | 1.44 [0.97-2.12] | 1.26 [0.81-1.95] |
|  | Motor delay | 41 (29.7%) | 7,606 (18.8%) | | 1.48 [1.01-2.17] | 1.31 [0.84-2.02] |
|  | Difficulty gaining weight | 11 (8.0%) | 1,625 (4.0%) | | 1.36 [0.74-2.52] | 1.19 [0.59-2.40] |
|  | Short stature | 5 (3.6%) | 892 (2.2%) | | NA | NA |
|  | Microcephaly | 6 (4.3%) | 426 (1.1%) | | NA | NA |
|  | Macrocephaly | 6 (4.3%) | 1,303 (3.2%) | | NA | NA |
|  | Obesity | 1 (0.7%) | 753 (1.9%) | | NA | NA |
|  | Other growth condition | 4 (2.9%) | 803 (2.0%) | | NA | NA |
|  | Encopresis | 20 (14.5%) | 3,464 (8.6%) | | 1.60 [0.97-2.61] | 1.05 [0.45-2.45] |
|  | Enuresis | 25 (18.1%) | 3,698 (9.1%) | | 1.75 [1.11-2.76] | 1.66 [0.83-3.34] |
|  | Sleep Disorder | 39 (28.3%) | 8,400 (20.8%) | | 1.23 [0.83-1.82] | 1.01 [0.64-1.58] |
|  | Strabismus | 10 (7.2%) | 1,282 (3.2%) | | 1.48 [0.76-2.88] | 1.36 [0.72-2.60] |
|  | Deafness/hearing loss | 11 (8.0%) | 803 (2.0%) | | 3.48 [1.81-6.68] | 3.35 [1.76-6.37] |
|  | Seizure disorder or epilepsy | 31 (22.5%) | 2,093 (5.2%) | | 3.23 [2.00-5.21] | 3.23 [2.00-5.21] |

* Percentages are column percentages
Model 1: The model for each comorbidity was adjusted for maternal and paternal education and age at delivery, child’s gender and year of birth, age of the child at evaluation, survey version, race, and annual household income.

Model 2: In addition to the covariates above, the model for each comorbidity was adjusted for other comorbidities with the same cluster (outcomes in the same cluster are highlighted in the same color)

Table S8 – Associations between infection in pregnancy and comorbidity in individuals with ASD

| **Exposure** |  | **Exposed (n = 115)** | | **Unexposed (n = 40,467)** | **Model 1** | **Model 2** |
| --- | --- | --- | --- | --- | --- | --- |
|  | **Outcome** | **n (%)** | | **n (%)** | **OR [95% CI]** | **OR [95% CI]** |
| Infection in pregnancy | Anxiety disorder | 31 (27.0%) | | 7,228 (17.9%) | 1.16 [0.73-1.83] | 0.96 [0.56-1.67] |
|  | Separation Anxiety | 16 (13.9%) | 2,265 (5.6%) | | 1.65 [0.92-2.97] | 1.35 [0.60-3.05] |
|  | Social Anxiety/Social Phobia | 19 (16.5%) | 3,589 (8.9%) | | 1.36 [0.79-2.34] | 1.19 [0.64-2.20] |
|  | Bipolar Disorder | 5 (4.3%) | 688 (1.7%) | | NA | NA |
|  | Depression or dysthymia | 12 (10.4%) | 2,491 (6.2%) | | 1.26 [0.66-2.41] | 1.14 [0.54-2.40] |
|  | Disruptive Mood Dysregulation Disorder | 8 (7.0%) | 1,278 (3.2%) | | NA | NA |
|  | Hoarding | 1 (0.9%) | 594 (1.5%) | | NA | NA |
|  | Obsessive-Compulsive Disorder | 15 (13.0%) | 3,186 (7.9%) | | 1.17 [0.65-2.09] | 1.00 [0.51-1.95] |
|  | ADHD or ADD | 56 (48.7%) | 14,251 (35.2%) | | 1.13 [0.75-1.72] | 0.97 [0.63-1.49] |
|  | Conduct Disorder | 11 (9.6%) | 844 (2.1%) | | 3.27 [1.55-6.93] | 2.92 [1.34-6.38] |
|  | Oppositional Defiant Disorder | 18 (15.7%) | 3,026 (7.5%) | | 1.45 [0.83-2.53] | 1.12 [0.62-2.03] |
|  | Tourette Syndrome or Tic Disorder | 9 (7.8%) | 1,118 (2.8%) | | NA | NA |
|  | Intellectual disability | 43 (37.4%) | 8,753 (21.6%) | | 1.63 [1.12-2.39] | 1.43 [0.92-2.21] |
|  | Learning disability | 45 (39.1%) | 9,496 (23.5%) | | 1.47 [0.98-2.22] | 1.18 [0.74-1.88] |
|  | Social Communication Disorder | 37 (32.2%) | 8,009 (19.8%) | | 1.60 [1.07-2.40] | 1.40 [0.90-2.19] |
|  | Motor delay | 38 (33.0%) | 7,609 (18.8%) | | 1.34 [0.88-2.03] | 1.04 [0.66-1.64] |
|  | Difficulty gaining weight | 21 (18.3%) | 1,615 (4.0%) | | 2.55 [1.54-4.20] | 2.95 [1.71-5.08] |
|  | Short stature | 7 (6.1%) | 890 (2.2%) | | NA | NA |
|  | Microcephaly | 4 (3.5%) | 428 (1.1%) | | NA | NA |
|  | Macrocephaly | 7 (6.1%) | 1,302 (3.2%) | | NA | NA |
|  | Obesity | 3 (2.6%) | 751 (1.9%) | | NA | NA |
|  | Other growth condition | 4 (3.5%) | 803 (2.0%) | | NA | NA |
|  | Encopresis | 16 (13.9%) | 3,468 (8.6%) | | 1.05 [0.60-1.83] | 0.66 [0.29-1.48] |
|  | Enuresis | 22 (19.1%) | 3,701 (9.1%) | | 1.45 [0.89-2.37] | 1.90 [0.96-3.78] |
|  | Sleep Disorder | 36 (31.3%) | 8,403 (20.8%) | | 1.18 [0.78-1.79] | 1.07 [0.65-1.77] |
|  | Strabismus | 10 (8.7%) | 1,282 (3.2%) | | 1.57 [0.79-3.13] | 1.38 [0.70-2.70] |
|  | Deafness/hearing loss | 11 (9.6%) | 803 (2.0%) | | 3.35 [1.72-6.53] | 3.30 [1.72-6.32] |
|  | Seizure disorder or epilepsy | 15 (13.0%) | 2,109 (5.2%) | | 1.53 [0.90-2.59] | 1.53 [0.90-2.59] |

* Percentages are column percentages
Model 1: The model for each comorbidity was adjusted for maternal and paternal education and age at delivery, child’s gender and year of birth, age of the child at evaluation, survey version, race, and annual household income.

Model 2: In addition to the covariates above, the model for each comorbidity was adjusted for other comorbidities with the same cluster (outcomes in the same cluster are highlighted in the same color)

Table S9 – Associations between lead poisoning and comorbidity in individuals with ASD

| **Exposure** |  | **Exposed (n = 130)** | | **Unexposed (n = 40,452)** | **Model 1** | **Model 2** |
| --- | --- | --- | --- | --- | --- | --- |
|  | **Outcome** | **n (%)** | | **n (%)** | **OR [95% CI]** | **OR [95% CI]** |
| Lead poisoning | Anxiety disorder | 34 (26.2%) | | 7,225 (17.9%) | 1.45 [0.94-2.24] | 1.24 [0.75-2.04] |
|  | Separation Anxiety | 14 (10.8%) | 2,267 (5.6%) | | 1.37 [0.75-2.52] | 0.84 [0.40-1.78] |
|  | Social Anxiety/Social Phobia | 20 (15.4%) | 3,588 (8.9%) | | 1.34 [0.81-2.21] | 0.97 [0.52-1.81] |
|  | Bipolar Disorder | 4 (3.1%) | 689 (1.7%) | | NA | NA |
|  | Depression or dysthymia | 15 (11.5%) | 2,488 (6.2%) | | 1.43 [0.81-2.51] | 1.11 [0.58-2.14] |
|  | Disruptive Mood Dysregulation Disorder | 9 (6.9%) | 1,277 (3.2%) | | NA | NA |
|  | Hoarding | 2 (1.5%) | 593 (1.5%) | | NA | NA |
|  | Obsessive-Compulsive Disorder | 20 (15.4%) | 3,181 (7.9%) | | 1.65 [1.00-2.72] | 1.43 [0.77-2.63] |
|  | ADHD or ADD | 59 (45.4%) | 14,248 (35.2%) | | 1.22 [0.82-1.80] | 1.11 [0.72-1.70] |
|  | Conduct Disorder | 5 (3.8%) | 850 (2.1%) | | NA | NA |
|  | Oppositional Defiant Disorder | 17 (13.1%) | 3,027 (7.5%) | | 1.26 [0.73-2.19] | 1.11 [0.58-2.13] |
|  | Tourette Syndrome or Tic Disorder | 10 (7.7%) | 1,117 (2.8%) | | 2.06 [1.03-4.12] | 2.00 [0.98-4.06] |
|  | Intellectual disability | 51 (39.2%) | 8,745 (21.6%) | | 2.14 [1.48-3.08] | 1.82 [1.20-2.76] |
|  | Learning disability | 56 (43.1%) | 9,485 (23.4%) | | 1.95 [1.33-2.86] | 1.57 [1.04-2.38] |
|  | Social Communication Disorder | 35 (26.9%) | 8,011 (19.8%) | | 1.46 [0.99-2.16] | 1.14 [0.76-1.73] |
|  | Motor delay | 33 (25.4%) | 7,614 (18.8%) | | 1.32 [0.86-2.01] | 0.93 [0.59-1.47] |
|  | Difficulty gaining weight | 9 (6.9%) | 1,627 (4.0%) | | NA | NA |
|  | Short stature | 6 (4.6%) | 891 (2.2%) | | NA | NA |
|  | Microcephaly | 2 (1.5%) | 430 (1.1%) | | NA | NA |
|  | Macrocephaly | 8 (6.2%) | 1,301 (3.2%) | | NA | NA |
|  | Obesity | 12 (9.2%) | 742 (1.8%) | | 4.07 [2.15-7.73] | 3.98 [2.07-7.64] |
|  | Other growth condition | 6 (4.6%) | 801 (2.0%) | | NA | NA |
|  | Encopresis | 16 (12.3%) | 3,468 (8.6%) | | 1.26 [0.73-2.16] | 0.73 [0.36-1.47] |
|  | Enuresis | 21 (16.2%) | 3,702 (9.2%) | | 1.57 [0.96-2.57] | 1.21 [0.62-2.39] |
|  | Sleep Disorder | 53 (40.8%) | 8,386 (20.7%) | | 2.24 [1.56-3.22] | 2.25 [1.54-3.27] |
|  | Strabismus | 8 (6.2%) | 1,284 (3.2%) | | NA | NA |
|  | Deafness/hearing loss | 5 (3.8%) | 809 (2.0%) | | NA | NA |
|  | Seizure disorder or epilepsy | 15 (11.5%) | 2,109 (5.2%) | | 1.92 [1.06-3.50] | 1.92 [1.06-3.50] |

* Percentages are column percentages
Model 1: The model for each comorbidity was adjusted for maternal and paternal education and age at delivery, child’s gender and year of birth, age of the child at evaluation, survey version, race, and annual household income.

Model 2: In addition to the covariates above, the model for each comorbidity was adjusted for other comorbidities with the same cluster (outcomes in the same cluster are highlighted in the same color)

Table S10 – Comparison of results from imputed dataset vs complete case analysis (sensitivity analysis) on associations between preterm birth and comorbidity in individuals with ASD

| **Exposure** |  | **Full dataset with imputed missing covariates, n = 40,582** | **Complete case analysis,**  **n = 24,365** |
| --- | --- | --- | --- |
|  | **Outcome** | **OR [95% CI]** | **OR [95% CI]** |
| Preterm birth | Anxiety disorder | 1.30 [1.20-1.42] | 1.32 [1.18-1.47] |
|  | Separation Anxiety | 1.61 [1.42-1.81] | 1.64 [1.40-1.92] |
|  | Social Anxiety/Social Phobia | 1.29 [1.16-1.44] | 1.34 [1.17-1.54] |
|  | Bipolar Disorder | 1.18 [0.94-1.48] | 1.07 [0.79-1.45] |
|  | Depression or dysthymia | 1.02 [0.89-1.17] | 1.03 [0.87-1.22] |
|  | Disruptive Mood Dysregulation Disorder | 1.12 [0.95-1.33] | 1.10 [0.88-1.37] |
|  | Hoarding | 1.59 [1.27-1.98] | 1.72 [1.30-2.27] |
|  | Obsessive-Compulsive Disorder | 1.25 [1.12-1.39] | 1.25 [1.08-1.44] |
|  | ADHD or ADD | 1.21 [1.12-1.30] | 1.21 [1.10-1.33] |
|  | Conduct Disorder | 0.81 [0.65-1.02] | 0.86 [0.64-1.17] |
|  | Oppositional Defiant Disorder | 1.20 [1.07-1.35] | 1.12 [0.97-1.31] |
|  | Tourette Syndrome or Tic Disorder | 1.12 [0.93-1.35] | 1.15 [0.90-1.46] |
|  | Intellectual disability | 1.22 [1.14-1.32] | 1.27 [1.15-1.40] |
|  | Learning disability | 1.21 [1.13-1.31] | 1.22 [1.10-1.34] |
|  | Social Communication Disorder | 1.12 [1.03-1.21] | 1.10 [1.00-1.22] |
|  | Motor delay | 1.59 [1.47-1.71] | 1.65 [1.49-1.81] |
|  | Difficulty gaining weight | 2.38 [2.09-2.71] | 2.22 [1.88-2.64] |
|  | Short stature | 2.05 [1.71-2.46] | 1.98 [1.55-2.53] |
|  | Microcephaly | 1.92 [1.47-2.50] | 1.96 [1.36-2.83] |
|  | Macrocephaly | 1.48 [1.26-1.74] | 1.51 [1.24-1.85] |
|  | Obesity | 1.44 [1.17-1.78] | 1.56 [1.18-2.08] |
|  | Other growth condition | 1.42 [1.17-1.72] | 1.45 [1.11-1.88] |
|  | Encopresis | 1.26 [1.13-1.40] | 1.23 [1.07-1.42] |
|  | Enuresis | 1.28 [1.15-1.41] | 1.25 [1.09-1.43] |
|  | Sleep Disorder | 1.28 [1.18-1.38] | 1.33 [1.20-1.46] |
|  | Strabismus | 1.47 [1.27-1.71] | 1.42 [1.17-1.73] |
|  | Deafness/hearing loss | 1.48 [1.22-1.80] | 1.52 [1.18-1.96] |
|  | Seizure disorder or epilepsy | 1.25 [1.09-1.42] | 1.18 [0.99-1.41] |

Note: The model for each comorbidity was adjusted for maternal and paternal education and age at delivery, child’s gender and year of birth, age of the child at evaluation, survey version, race, and annual household income.

Table S11 – Comparison of results from imputed dataset vs complete case analysis (sensitivity analysis) on associations between hypoxia at birth and comorbidity in individuals with ASD

| **Exposure** |  | **Full dataset with imputed missing covariates, n = 40,582** | **Complete case analysis,**  **n = 24,365** |
| --- | --- | --- | --- |
|  | **Outcome** | **OR [95% CI]** | **OR [95% CI]** |
| Hypoxia at birth | Anxiety disorder | 1.16 [1.04-1.29] | 1.13 [0.98-1.30] |
|  | Separation Anxiety | 1.23 [1.05-1.44] | 1.10 [0.89-1.36] |
|  | Social Anxiety/Social Phobia | 1.27 [1.11-1.46] | 1.23 [1.02-1.47] |
|  | Bipolar Disorder | 1.10 [0.82-1.48] | 1.06 [0.71-1.59] |
|  | Depression or dysthymia | 1.28 [1.08-1.51] | 1.30 [1.05-1.60] |
|  | Disruptive Mood Dysregulation Disorder | 1.43 [1.16-1.75] | 1.44 [1.10-1.87] |
|  | Hoarding | 1.21 [0.89-1.67] | 1.23 [0.81-1.88] |
|  | Obsessive-Compulsive Disorder | 1.25 [1.09-1.45] | 1.14 [0.94-1.37] |
|  | ADHD or ADD | 1.23 [1.12-1.35] | 1.12 [0.98-1.27] |
|  | Conduct Disorder | 1.21 [0.91-1.62] | 1.37 [0.93-2.01] |
|  | Oppositional Defiant Disorder | 1.12 [0.96-1.30] | 1.08 [0.88-1.31] |
|  | Tourette Syndrome or Tic Disorder | 1.16 [0.92-1.47] | 1.00 [0.71-1.40] |
|  | Intellectual disability | 1.46 [1.33-1.61] | 1.39 [1.23-1.58] |
|  | Learning disability | 1.44 [1.31-1.58] | 1.37 [1.21-1.55] |
|  | Social Communication Disorder | 1.40 [1.27-1.55] | 1.37 [1.20-1.56] |
|  | Motor delay | 1.76 [1.60-1.94] | 1.70 [1.50-1.92] |
|  | Difficulty gaining weight | 1.78 [1.52-2.08] | 1.74 [1.41-2.14] |
|  | Short stature | 1.76 [1.42-2.19] | 1.80 [1.35-2.42] |
|  | Microcephaly | 1.43 [1.04-1.97] | 1.53 [0.99-2.35] |
|  | Macrocephaly | 1.54 [1.27-1.88] | 1.76 [1.37-2.24] |
|  | Obesity | 1.58 [1.23-2.03] | 1.57 [1.11-2.22] |
|  | Other growth condition | 1.56 [1.22-2.00] | 1.53 [1.09-2.12] |
|  | Encopresis | 1.51 [1.32-1.71] | 1.61 [1.36-1.90] |
|  | Enuresis | 1.45 [1.28-1.65] | 1.44 [1.22-1.70] |
|  | Sleep Disorder | 1.45 [1.31-1.59] | 1.39 [1.22-1.58] |
|  | Strabismus | 1.66 [1.38-2.00] | 1.67 [1.32-2.12] |
|  | Deafness/hearing loss | 1.72 [1.37-2.16] | 1.74 [1.29-2.34] |
|  | Seizure disorder or epilepsy | 1.83 [1.57-2.13] | 1.77 [1.43-2.18] |

Note: The model for each comorbidity was adjusted for maternal and paternal education and age at delivery, child’s gender and year of birth, age of the child at evaluation, survey version, race, and annual household income.
